# Decision Considerations and Strategies for Lip Surgery in Patients with Cleft lip/Palate: A Qualitative Study

**DOI:** 10.1101/2023.04.20.23287416

**Authors:** Carroll Ann Trotman, Julian Faraway, M. Elizabeth Bennett, G. David Garson, Ceib Phillips, Richard Bruun, Renie Daniel, Lisa Renee David, Ingrid Ganske, Lauren K Leeper, Carolyn R. Rogers-Vizena, Christopher Runyan, Andrew R. Scott, Jeyhan Wood

**Affiliations:** College of Dentistry, The Ohio State University, Ohio, United States of America; Department of Mathematical Sciences, University of Bath, Bath, United Kingdom; Department of Orthodontics, UNC Adams School of Dentistry, Chapel Hill, North Carolina; Department of Public Administration, School of Public and International Affairs, North Carolina State University, Raleigh, North Carolina; Department of Developmental Biology, Harvard School of Dental Medicine; Department of Oral and Maxillofacial Surgery, UNC Adams School of Dentistry, Chapel Hill, North Carolina; Department of Plastic and Reconstructive Surgery, Wake Forest Baptist Health, Wake Forest, North Carolina; Department of Plastic & Oral Surgery, Boston Children’s Hospital, Boston, Massachusetts; Department of Otolaryngology/Head and Neck Surgery, University of North Carolina Health, Chapel Hill, North Carolina; Department of Plastic & Oral Surgery Boston Children’s Hospital, Boston, Massachusetts; Department of Plastic Surgery, Wake Forest Baptist Health, Wake Forest, North Carolina; Department of Otolaryngology, Tufts University School of Medicine, Boston, Massachusetts; Department of Plastic Surgery, University of North Carolina Health, Chapel Hill, North Carolina

**Keywords:** Cleft lip/palate decision-making clinical trial lip repair lip revision qualitative

## Abstract

**Objective:** To qualitatively assess surgeons decision making for lip surgery in patients with cleft lip/palate (CL/P).

**Design:** Prospective, non-randomized, clinical trial.

**Setting:** Clinical data institutional laboratory setting.

**Patients, Participants:** The study included both patient and surgeon participants recruited from four craniofacial centers. The patient participants were babies with a CL/P requiring primary lip repair surgery (n=16) and adolescents with repaired CL/P who may require secondary lip revision surgery (n=32). The surgeon participants (n=8) were experienced in cleft care. Facial imaging data that included 2D images, 3D images, videos, and objective 3D visual modelling of facial movements were collected from each patient, and compiled as a collage termed the Standardized Assessment for Facial Surgery (SAFS) for systematic viewing by the surgeons.

**Interventions:** The SAFS served as the intervention. Each surgeon viewed the SAFS for six distinct patients (two babies and four adolescents) and provided a list of surgical problems and goals. Then an in-depth-interview (IDI) was conducted with each surgeon to explore their decision-making processes. IDIs were conducted either in person or virtually, recorded, and then transcribed for qualitative statistical analyses using the Grounded Theory Method.

**Results:** Rich narratives/themes emerged that included timing of the surgery; risks/limitations and benefits of surgery; patient/family goals; planning for muscle repair and scarring; multiplicity of surgeries and their impact; and availability of resources. For diagnoses/treatments, surgeons agreed, and level of surgical experience was not a factor.

**Conclusions:** The themes provided important information to populate a checklist of considerations to serve as a guide for clinicians.

## Introduction

For patients with cleft lip/palate (CL/P) there exists little information on surgeons’ decision-making process for primary lip repair and secondary lip revision surgery. These decisions affect the post-surgical esthetic and functional outcomes of the nasolabial region. The condition itself occurs once in every 600 to 800 births, and patients have their initial or primary lip repair soon after birth. The impact of surgery on facial soft tissue form and movement is highly variable.^1^ For many, soft tissue disabilities persist in the form of facial disfigurements and impaired facial and circumoral movements that often require additional revision surgeries.^2–3^ The burden of care is great and includes (1) Direct costs for treatment expenses, ancillary services, and time lost at work as well as indirect costs such as the health/emotional well-being of the child and caregivers, and these costs have increased over time.^4^ (2) Reports by parents that the quality of parent-infant interactions is adversely affected;^5^ that their children develop psychological problems because of their facial appearance,^5^ and later in life many wish to have additional surgery.^6^ (3) Patient complaints of anxiety and awkward moments because of their facial stigmata during social interactions with other non-stigmatized individuals.^7^ (4) Evidence that less successful surgeries may impact patients socioeconomically in the form of diminished income and educational accomplishments compared with their non-cleft counterparts.^8–9^

The decision to perform a lip revision surgery relies mainly on a subjective assessment of the patient’s facial structure or form made by the surgeon in conjunction with the goals of the patient/caregivers. The important role of movement has been given less consideration mainly because of challenges to assess and improve movement (e.g., the amount/quality of the tissue available). Even when surgeons attempt to assess movement, there are no quantitative/visual aids to incorporate movement into treatment planning decisions. To that end, our research group has developed a systematic assessment approach termed the *Standardized Assessment for Facial Surgery or SAFS* that is used to quantify facial disability for surgical treatment planning and outcome assessment purposes.^10–12^ It incorporates a unique collage of three-dimensional (3D) facial quantitative dynamic and static measures and visual dynamic comparisons of patients soft tissue movements versus controls for an objective assessment of patients’ faces and facial movements, as well as data comprising 2D, 3D, and video facial images for a subjective assessment. The approach allows surgeons to broaden their “vista” of a patient’s problems by identifying ‘movement’ as well as ‘form’ problems. They then have the potential to make decisions that better address problems on a patient specific basis.^3.12^

When using the SAFS to aid in treatment planning, surgeons are required to mentally integrate multiple sources of information/data^13, 14^ along a continuum ranging from intuitive and subconscious (SAFS subjective data) to analytical and conscious (SAFS objective data).

Decisions are reached by a combination of each according to the complexity of the situation and the experience of the surgeon. Thus, given that these types of ‘expert decisions’ are a relatively unexplored area in the surgical sciences, the *primary* objective of this study was to assess how surgeons integrate the SAFS objective measures and visual aids with the SAFS systematic subjective assessment in decision-making for lip surgery in patients with CL/P. This study tests the hypothesis that when surgeons are presented with individual patients’ SAFS data, common themes emerge from their decisions for the surgical management of children with repaired CL/P who may potentially benefit from revision surgery and infants with unrepaired CL/P requiring initial lip repair. Secondary areas explored were whether surgeons agree on the diagnosis and treatment planning for lip revision and on the nuances of the surgery for lip repair; and whether surgical experience in terms of years spent treating patients with CL/P affect the decision-making process.

## Method

This research was funded by a grant from the National Institutes of Health NIDCR branch (Grant # U01 DE024503). The participants for this qualitative study were part of a non-randomized clinical trial (NCT03537976) conducted at six Craniofacial Centers that served as recruitment sites for the patient and surgeon participants. The centers were The University of North Carolina (UNC) and Wake Forest Baptist Health Craniofacial Centers in Chapel Hill and Winston Salem, North Carolina; and Boston Children’s Hospital, Massachusetts General Hospital, Shriners Hospitals for Children, and Tufts Medical Center in Boston, Massachusetts. Tufts University School of Dental Medicine (TUSDM) Boston, Massachusetts served as the data coordinating center. The study protocol (see supporting information) and consent and HIPAA documents were approved by the Tufts Health Sciences and UNC Biomedical Human Subjects Institutional Review Boards. The authors declare no conflict of interest in preparing this article.

### Patient Participation

At each Center, the patient’s surgeon made the initial clinical decision to perform either primary lip repair or revision surgery. Once this decision was made, the patients’ medical history was screened to determine eligibility (Table 1). If eligible, patients were recruited to participate, and data collection visit(s) scheduled. Patients were enrolled in two groups. Group 1 (age range 4 to 21 years) comprised two sub-groups: Patients who were recommended for, and could benefit from, lip revision surgery (n=22, termed revision), and patients who were not recommended for revision (n=10, termed non-revision). The non-revision sub-group served as a negative control for the revision sub-group. The addition of the non-revision subgroup was an IRB approved modification to the original protocol. Group 2 (age range birth to 8 months) were infants scheduled for primary lip repair (n=16, termed repair).

**Table 1.** Revision and non-revision patients, lip repair infants, and surgeon eligibility criteria.

| **Participants** | **Inclusion Criteria** | **Exclusion Criteria** |
| --- | --- | --- |
| Revision & Non-Revision | • For the revision patients, presence of a previously repaired unilateral or bilateral cleft lip and palate with a complete cleft of the primary palate and at least a partial or complete cleft of the secondary palate.<br>• For the revision patients, a professional clinical recommendation by the craniofacial plastic / oral maxillofacial surgeon for a full or partial thickness lip revision.<br>• For the revision and non-revision patients, patient (depending on age) and guardian have ability to comprehend verbal instructions in English, Spanish, or Chinese.<br>• For the revision and non-revision patients, patient (depending on age) or guardian able to give consent / assent and have an ability to provide a signed and dated informed consent form in English, Spanish, or Chinese.<br>• For the revision and non-revision patients, patient (depending on age) or guardian willing to comply with all study procedures and be available for up to 2 study visits.<br>• For the revision and non-revision patients, patient age 4 to 21 years. | • Lip revision surgery within the past year.<br>• A diagnosis of a craniofacial anomaly other than cleft lip (and palate).<br>• A medical history of collagen vascular disease, or systemic neurologic impairment.<br>• Mental, visual, or hearing impairment to the extent that comprehension or ability to perform tests associated with the collection of the imaging data is hampered. |
| Repair | • Patient has presence of an unrepaired unilateral or bilateral cleft lip and palate with a complete cleft of the primary palate and at least a partial or complete cleft of the secondary palate.<br>• Parent/guardian is willing and able to provide a signed and dated informed consent form in English, Spanish, and Chinese.<br>• Parent/guardian is willing and able to comprehend verbal instructions in English, Spanish, or Chinese.<br>• Parent/guardian is willing and able to comply with all study procedures and be available for the duration of the study visits.<br>• Patient age birth to 8 months. | • A diagnosis of a craniofacial anomaly other than cleft lip (and palate).<br>• A medical diagnosis of collagen vascular disease, and systemic neurologic impairment.<br>• Mental, visual, or hearing impairment to the extent that the infant's ability to perform tests associated with the collection of the imaging data is hampered. |
| Surgeon | • Surgeons experienced in the care and treatment of patients with CL/P.<br>• Surgeons should belong to a Cleft lip and palate team as defined by the ACPA Team standards document (www.acpa-cpf.org/team_care/).<br>• Surgeon should have a minimum of 2 lip revision and 2 lip repair surgeries per 12-month period.<br>• The total number of surgeons represents a broad range of experience specific to the length of time the surgeon has been treating patients with cleft lip and palate and the length of service on | • Surgeons who have previously used the 3D facial images and the dynamic facial movement data of the SAFS protocol prior to their enrollment in this study. |
|  | a cleft lip and palate team. Ideally, the length of time specific to surgical experience will include the following three categories: Surgeon-raters with < 5 years of experience and $\geq 5$ years of experience.<br>• Surgeons' willingness to participate and availability for the duration of the study. |  |

Figure 1 is a schematic of the trial design and logistics. The patients attended Facial Animation Laboratories located at TUSDM and UNC for data collection visit(s). For the revision and repair patients, a first visit occurred at no greater than three months and up to one day before the scheduled surgery, and if needed a second visit occurred to complete data collection up to one day before surgery. For the non-revision patients, the first visit occurred at any time with a second visit up to one month after the first if needed. At the time of the first visit, written consent was obtained from the patient and/or the patients’ parent/carer, and the subjective and objective data for the SAFS were collected from the patients. All patients continued to receive other services routinely provided by their respective Center.

**Figure 1.**
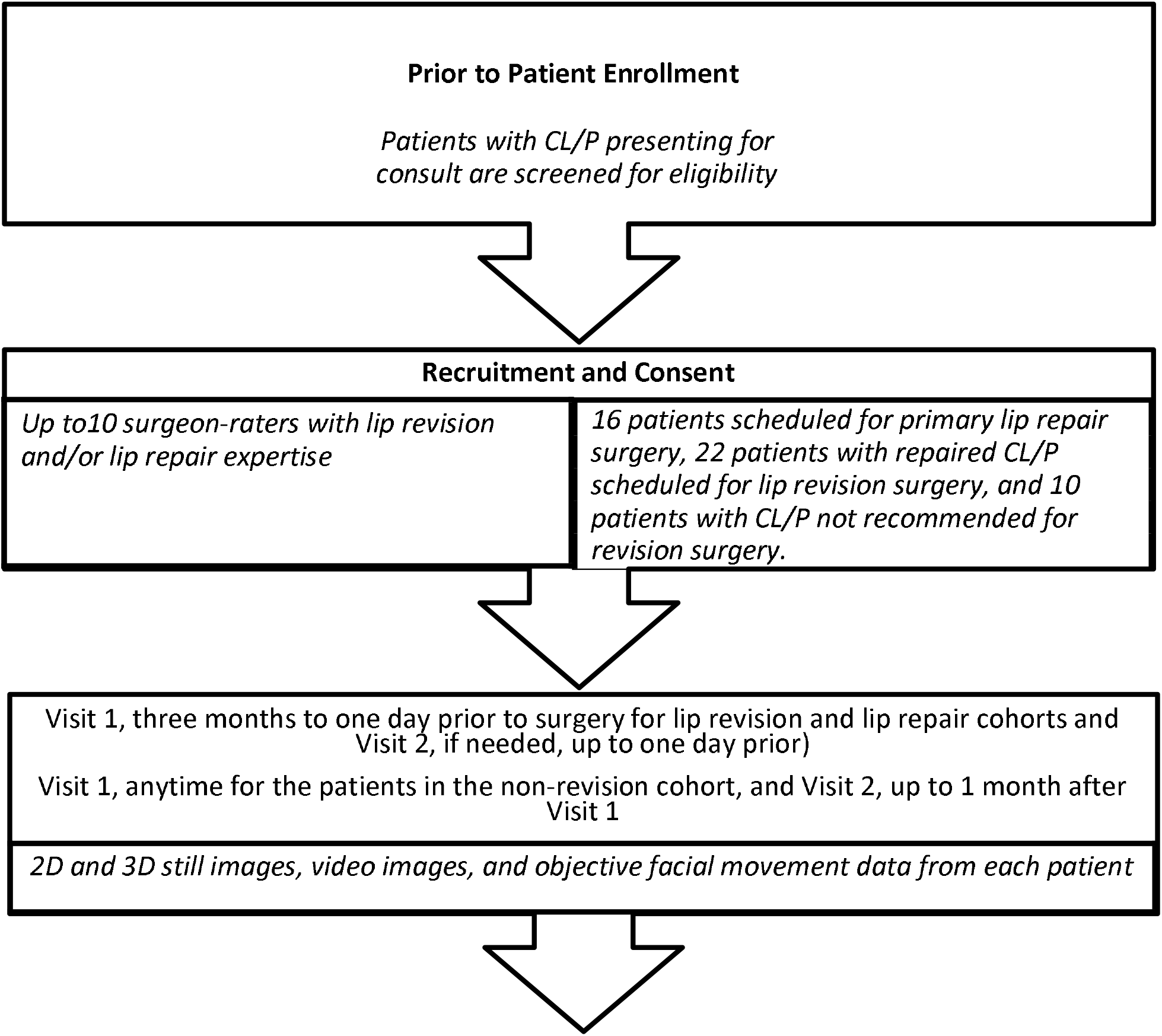
Schematic of the trial design and logistics.

### Surgeon Participation

Up to ten surgeons (and no less than eight surgeons) were previously recruited to participate in the trial based on the eligibility criteria (Figure 1, Table 1). The surgeons had high volume practices devoted to the care of patients with CL/P and were selected based on their different levels of surgical experience to obtain a broad range of feedback on diagnosis and treatment planning. Prior to participating in the trial, they were trained in the use of the SAFS, after which, they each completed the SAFS on six study patients: three revision, one non-revision, and two repair patients. Because in this study the mode of presentation of the patients’ data for clinical evaluation by the surgeons was via the SAFS, it was deemed important by the medical monitor that for patient safety, surgeons would not evaluate the SAFS on their own patients. Therefore, the surgeons completed the SAFS on patients treated at Centers other than their own, and for patients in Group 1, the surgeons were unaware of whether a patient was in the revision or non-revision sub-groups, that is, whether a patient was recommended for revision.

Additionally, to assess agreement among the surgeons with the use of the SAFS, the SAFS of two patients in each revision and repair group were repeated with the surgeons. This was a protocol modification approved by the IRB.

### SAFS Presentation and Surgeons’ Interviews

The SAFS standardized facial images of a patient were presented in four sequential Phases. The following demonstrates these phases for a revision patient. Phase 1. 2D facial photographs at rest and at the maximum of different animations https://tufts.box.com/s/c8yf5ot57siyg3odre0un6u67tc3ng66. Phase 2. 3D facial photographs at rest and at the maximum of different animations https://tufts.box.com/s/77xbh3pgsl9svl4q1l20beyanhaapwyu. Phase 3. Video recordings of the face during different animations https://tufts.box.com/s/vjyfcz2m3p86qd8lbimnrb88vxuyi0lj. Phase 4. Dynamic objective measures displayed as visual aids of the face during different animations https://tufts.box.com/s/5436b628hydgc7zja79v9qrrlpp7b22w and vector plots of movements during different animations https://tufts.box.com/s/rlk67gy7rye66tg4bpnwbe1u4f92gfvf. One researcher (CAT) presented the SAFS to each surgeon independently. Following each phase, the surgeon provided a detailed problem list and goals for the patient’s surgery. Then, a 45 to 60 minute, semi-structured, one-on-one, in-depth-interview (IDI) was conducted via phone with each surgeon to qualitatively explore their diagnosis/treatment planning and decision-making process. The interviewer was a clinical psychologist with experience and skill in focused discussions in both corporate research and academic settings. Interview guides were utilized that were inclusive and designed to elicit open-ended discussion. Initially, areas explored were modeled on those identified by surgeons involved in the development of the SAFS and by surgeons in the current trial during their training in the use of the SAFS. As is the practice in this type of qualitative research, the guides were modified during the project period based on surgeons’ responses. All the SAFS transcripts including the presentations and interviews were audiotaped, transcribed, and stripped of identifying data.

### Data Analyses

The IDI transcripts were analyzed using the Grounded Theory Method (GTM).^15–17^ Under GTM, themes are induced from open-ended responses rather than from *a priori* conceptual categories. Making sense of complex interview data via GTM mitigates researcher bias and supports openness to results not expected beforehand,^18^ as under Burawoy’s “extended case method”.^19–21^ ATLAS.ti statistical software^22–23^ was used to analyze the data that included labeling, categorizing, and sorting the large amounts of interview data tagged through open coding. Codes then were iteratively associated with core code categories based on centrality to a code group topic. Based on the method of constant comparison, stopping occurred at the point of theoretical saturation. Topical reports then were generated based on the surgeons’ quotations collected in code groups.

### Sample Size

Sample size in qualitative research is set at the point when the data collected reveals no new themes/concepts or patterns (theoretical saturation). There is no clear consensus on appropriate sample sizes; however, in this type of qualitative research evidence suggests that saturation generally occurs between 10 and 30 interviews.^15^ Also, while the GTM does not require an exact random sample of the population of surgeons specializing in cleft surgery, the sample in the present study can be said to represent a fair balance by such demographic factors as gender, age, and experience in cleft care.

## Results

The patients were recruited between January 2018 and 2020. The total number enrolled was 49 of which one was lost to follow-up—thus, 48 patients (Table 2) completed the SAFS data collection. Eight of the 10 surgeons who were recruited completed the study. The two surgeons that did not were both from the same center—they did not view any patient SAFS. Of the eight surgeons, six were female and two were male. At the start of the study, four surgeons had less than five years of experience in the treatment of patients with CL/P and the other four had greater than or equal to five years of experience. As stated previously, to assess agreement among surgeons, it was decided to repeat a subset of patients. This decision was made close to the end of patient recruitment and as a result 36 of the 48 patient SAFS (Table 2) were used to complete the surgeon interviews. The additional 12 patient SAFS provided a buffer whereby as far as possible surgeons’ SAFS presentations were balanced by unilateral and bilateral CL/P patients. In all, a total of 48 IDIs were completed by the surgeons.

**Table 2.** Patient Demographics

| **All Patients Enrolled with SAFS Data (n=48)** |  |  |  |  |  |  |
| --- | --- | --- | --- | --- | --- | --- |
|  | Age | Male | Female | Unilateral | Bilateral | Total |
| Surgery Type |  |  |  |  |  |  |
| Revision (yrs.) | 12.2±4.8 | 13 | 8 | 15 | 6 | 21 |
| Non-Revision (yrs.) | 14.0±2.7 | 5 | 5 | 5 | 5 | 10 |
| Repair (mos.) | 3.1±0.8 | 13 | 4 | 10 | 7 | 17 |
| Total |  | 31 | 17 | 30 | 18 | 48 |
| **Patients whose SAFS Data were used by Surgeons (n=36)** |  |  |  |  |  |  |
|  | Age | Male | Female | Unilateral | Bilateral | Total |
| Surgery Type |  |  |  |  |  |  |
| Revision (yrs.)* | 11.9±4.7 | 11 | 7 | 13 | 5 | 18 |
| Non-Revision (yrs.) | 13.7±3.0 | 4 | 4 | 4 | 4 | 8 |
| Repair (mos.)** | 3.1±0.8 | 7 | 3 | 8 | 2 | 10 |
| Total patients |  | 22 | 14 | 25 | 11 | 36 |

\* Two additional patients from the 48 revision patients were shown to the same surgeons.
\*\* Two additional patients from the 17 repair patients were shown to the same surgeons.

Critical constructs of the qualitative transcript analyses included patient type (infant and child), type of surgery (revision, non-revision, and repair), CL/P (unilateral and bilateral), surgeon agreement (repeated patients), and surgeon experience (< 5 years and ≥ 5 years at the start of the trial). The transcripts’ themes represent diversity by these constructs enabling comparison of the main construct of interest and the surgical decision-making processes of the surgeons. Because there is no objective measure for “importance” of a theme, the results are ordered by number of quotations associated with a theme—the theme with the greatest number of quotations is presented first and that with the least presented last. We leave it to the reader to decide if ‘number’ of quotations is a surrogate for ‘importance’. Also, for each theme a sampling of illustrative surgeon quotations is provided (tagged by transcript document number) to lend richness to the analysis. Common themes emerged from the IDIs when the surgeons were engaged in treatment planning for lip revision (Table 3) and for primary lip repair (Table 4) with the use of the SAFS. Based on these themes the null hypothesis as cited was rejected.

**Table 3.** Common themes recognized in surgeons’ decisions for lip revision surgery.

| *Common Themes (Lip Revision)* |
| --- |
| 1. When it comes to facial/lip revision surgery timing is everything<br>2. All procedures involve risks and limitations<br>3. The treatment plan should follow patient and family goals<br>4. Plan ahead for the maneuvers that will be done during surgery<br>5. Muscle repair is tricky and options are limited<br>6. Scarring is a major consideration in planning for surgery<br>7. Results of lip (& nasal) revision will be evident and beneficial from several aspects—<br>appearance, speech, and psychosocially<br>8. Multiple surgeries are needed<br>9. Orthodontics/alveolar bone grafting and orthognathic surgery affect revision surgical<br>planning<br>10. Treatments impacted by surgeons' resources<br>11. Other (less discussed themes)<br>- Realistic expectations must be set<br>- Post-operative family and patient cooperation is important<br>- Some diagnoses and treatments are fairly standard<br>- Surgeons differ in surgical procedures they recommend<br>- Asymmetry is a common problem that can be fixed with surgery<br>- Effects of surgery on breathing must be a primary concern<br>- Not everything requires surgery<br>- Some things are discovered intraoperatively<br>- Ethnicity matters |

**Table 4.** Common themes recognized in surgeons decisions for lip repair surgery.

| *Common Themes (Lip Repair)* |
| --- |
| 1. Use of nasal alveolar molding (NAM) has pros and cons and there are alternatives<br>2. Working closely with the family is important<br>3. Diversity among patients impacts diagnosis, planning, and outcomes<br>4. Soft tissue work has special considerations<br>5. Lip repair surgery has risks and limitations<br>6. Different surgeons take different approaches to treatment<br>7. Other (less discussed themes)<br>- Live infant examinations provide important tactile information<br>- Telehealth is limited compared with face to face patient examination |

### (1) Lip Revision Surgery

#### Theme 1. When It Comes To Lip Revision Surgery, Timing Is Everything

This theme takes various forms. By far the most numerous comments revolve around the importance of the best timing for surgery. There is an expectation that a sequence of multiple surgeries or treatment interventions will be required (e.g., sometimes multiple staged revisions, orthodontics, alveolar bone grafting, jaw surgery, etc.). Ideally it is better to wait until craniofacial growth is complete—at approximately 17 to 18 years in males and slightly earlier in females—before doing jaw surgery if it is needed and /or aggressive nasal surgery such as rhinoplasty to correct the nasal dorsum and/or nasal septal deviation (septorhinoplasty)—septal deviation causes breathing issues in patients. Jaw surgery is a maxillary advancement but in certain instances the mandible also may be set back depending on the patient’s facial structure. In addition, it is preferred that alveolar bone grafting and jaw surgery be completed before definitive nasal work/rhinoplasty. If nasal work is done in childhood, it is best limited to less aggressive procedures such as columella lengthening, nasal tip adjustments, and/or adjustments to the nasal sills for symmetry—work that has limited effects on the nasal cartilage which may still have growth potential in children. However, when a child has functional needs (e.g. breathing problems) and/or psychosocial issues (e.g., being teased, a burning desire for a correction) surgeons may consider doing a lip/nasal revision at or around the time of bone grafting (∼ 6 to 12 years) but even then, it is preferred to delay the revision until at least 6 months after alveolar bone grafting because of expected swelling as a result of the bone graft, as is the ideal preference to delay lip/nasal revision until after jaw surgery because of expected facial changes that occur that will impact the patient’s final facial appearance.

- “…… There are some things that can’t be done at this age [CHILDHOOD]. Primarily, that has to do with some of the nasal stuff …… The breathing issues that go along with the history of cleft lip and palate involve septal deviation and that is not something that I would thoroughly address at this age because the septum is the growth center for the nose and it undergoes a lot of changes during the teenage years and then it also undergoes a lot of changes if you do a maxillary advancement, if he ends up having an underbite develop as he gets bigger. The work that I would do on the nose at this age would just involve the tip of the nose and straightening just the caudal end to the very tip of the septum, but [THE PATIENT] will eventually, almost certainly, need some further work on the septum but that would be done after full facial growth.”
- “The bone grafting has some very specific timing when it needs to happen. Lip revisions can be done anywhere along the way, driven by functional issues or issues with peers, but not an arbitrary idea of perfection.”

#### Theme 2. All Procedures Involve Risks And Limitations

The second-most discussed theme revolves around the risks of the surgical procedures and the limits to what is surgically possible. The risk of patients needing multiple surgeries is mentioned most-commonly. Specifically, surgeons are aware of the short-term risks of multiple surgeries such as bleeding, infection, risk of a cartilage graft becoming necrotic, and they recognize long-term effects such as the negative impact on midfacial growth resulting in midfacial deficiency. Bilateral cleft lip revision is considered particularly risk prone. Scarring is considered a common risk for the lip and nose which may result in impaired movements, hypertrophic tissue in the lip, and a negative impact on nasal growth. Hypertrophic tissue is particularly a problem in patients with darker skin color. Another risk is the possible adverse impact on speech resulting from jaw surgery (midfacial advancement) in later adolescence leading to a possible worsening of hypernasality—patients should be informed of this possibility. Surgeons are mindful that relapse is always possible especially after jaw surgery, and therefore as far as possible, jaw surgery should be delayed until craniofacial growth has stabilized. Also, when things do go wrong it can be very difficult to pinpoint the reason (e.g., poor lip movement after surgery). Some surgeons express the view that surgically altering lip movement in a predictable manner is not possible. They recognize that surgical perfection is near impossible in this patient population and sometimes even an ‘improvement’ may not be possible. Stretching the limits of surgery, for example, aggressive surgery, may lead to dangers of surgical overreach.

- “Aside from the typical surgical risks of bleeding and infection and risk of anesthesia, I would tell them [THE FAMILY] the risk of having to repeat the surgery is certainly present. If it’s a cartilage graft that I’m taking, which I probably would, the risk of that graft becoming necrotic. The biggest risk, I would say, would be the fact that [THE PATIENT] would need a secondary procedure…… growth is, I don’t want to say unfavorable because growth is independent it’s not favorable or unfavorable, growth is a good thing in …… but if [THE PATIENT] happens to grow in a way where the surgical repair can’t keep up then that would probably be the biggest risk.”
- “I think the scar burden is overall good, [THE PATIENT] is never going to be able to not have visible scars.”

#### Theme 3. The Treatment Plan Should Follow Patient And Family Goals

The third-most discussed theme centered on the need to place the patient and family goals first. The surgical decisions should be guided by the patient and family as their needs are often subjective and family specific. If the patient/family do not perceive an issue with the lip and/or nose then in most instances no interventions are necessary, alternatively, if a problem is perceived then attempts are made to address it. Although improvement in appearance is ideal, patient and parent satisfaction with the outcomes of surgery is primary. Surgeons recognize the need to strive for family and patient consensus on treatment, but the patient concerns are major and drive the expected treatment. Involving the patient and family in treatment decisions should start early, and options for surgery should be discussed in a non-directive manner. Re-confirming the treatment with the patient and family immediately pre-operatively is advised.

- “My goals of surgery are …… completely guided by the patient. So, if [THE PATIENT] says “No, I like the way my nose looks.” Then we don’t fix [THE] nose Or if [THE PATIENT’S] like, “I don’t care about my lip. I really could care less. The only thing I want is to be able to breathe out of the left side of my nose.” Then …… we might be a little bit more aggressive with that nasal surgery.”

- “I actually do ask them in the pre-op area to go back over certain things and then I tend to re-discuss with the parent and the child depending on their age, obviously, the things that, again, I’m going to focus on today. Kind of, “Is this what you guys understand that we’re working on today?”

#### Theme 4. Plan Ahead For The Maneuvers That Will Be Done During Surgery

There is universal agreement among surgeons on the importance of pre-surgical planning. The surgeons provide several patient specific surgical planning summaries, though only two are quoted below, this theme was the fourth-most discussed in the transcripts. They note the importance of planning maneuvers such as manipulating and/or grafting nasal cartilage and augmenting soft tissue. While the patient is under anesthesia fine measurements can be made to address asymmetries, and surgical markings can be drawn to assist with maneuvers such as columella lengthening and adjusting the philtrum width.

- “Once they’re asleep, I just do some fine measurements to check, double-check for asymmetries and make note of those. Then I draw up my surgical markings, which for the surgery that I proposed doing, involves making an incision at the base of the nose, beneath the nostrils, beneath the alar, beneath the columella. Then, after I make that incision, I really dissect down deep through the muscular sling that goes around the lip. You essentially detach the upper lip from the nose and allow the nose to be manipulated freely. Then, essentially, I do a cinch suture, which involves grabbing the corners of the nose and tightening ……[SURGEON CONTINUES]”

#### Theme 5. Muscle Repair Is Tricky And Options Are Limited

Issues related to the musculature and muscle repair are the fifth-most discussed theme. In part, this reflects the fact that the SAFS dynamic technology used by surgeons brought attention to movement issues that may not be apparent in the 2D and 3D images. Surgeons feel that they can address static lip form such as, for example, increasing or decreasing lip height or width to improve symmetry or replacing scar tissue but are limited in their muscle repair techniques to address directionality of soft tissue movement and even at times to completely re-approximate the muscle tissues. The lack of muscle tissue can severely limit movements—for example in a patient with a bilateral CL/P there may be limited muscle in the prolabium that results in a “tight appearance of the upper lip” post surgically. In most instances, revision is feasible, but when it is not, there may be alternatives—e.g., use of Botox to enhance or balance lip symmetry and/or facial physical therapy with biofeedback guidance to enhance facial expressive behaviors.

- “There are patients where the muscle, the orbicularis oris muscle, the one that goes around the mouth and allows you to purse your lips, there are cases where clearly, that muscle is not in continuity, it’s not connected, so there are some kind of crude corrections we can do. We can correct a muscle that’s not properly connected, but I don’t really have ways I know of addressing or of modifying the muscle to change horizontal lip movement directly or vertical lip movement or compaction. Other than just removing a complete portion of the lip in one direction or the other, but again, those are kind of-- generally I think of as more of a static change. We’re going to decrease the height or decrease the width.”

#### Theme 6. Scarring Is A Major Consideration In Planning For Surgery

Scarring is not just a risk factor for surgery, it is also a major treatment issue. Treatment of scarring is the sixth-most extensively discussed theme in the study. Scar revision surgery *may* improve appearance, but past scarring in a patient usually predicts future scarring, and there is a trade-off between scar removal (e.g., with a full thickness lip revision) and leaving sufficient tissue for movement. Moreover, when a patient has had multiple lip revisions, surgeons are more cautious when contemplating doing another due to the greater difficulty in getting a successful result. Thus, revision to remove scar tissue is not for everyone as described in the quote below. Non-surgical options to minimize scarring that include fat and /or Botox injections into the lip are mentioned.

- “I think [THE PATIENT] has a very favorable scarring for [THE] skin type, so as a surgeon I don’t want to go anywhere near it. You run the risk of taking someone with almost no scar and in your revision, giving them like a horrible scar ……”
- “We can …try to minimize those scars. We can do other things—the injection of fat underneath can help. We could do Botox underneath……these things will hopefully help [THE PATIENT] to have a positive result.”

*Themes 7-10* are summarized briefly below with illustrative surgeon quotations.

#### Theme 7. Results Of Lip (& Nasal) Revision Will Be Evident And Beneficial From Several Aspects—Appearance, Speech, and Psychosocially

- “[SURGEON REFERRING TO A GENERAL PATIENT] So, when she goes to the recovery area and everyone comes back to see her, they’re all going to be like “Wow. That doesn’t look like her at all.” Like “Who is this kid?” But they’re also would be like “Wow. She looks just like her sister now. [OR] She never looked like her mom, and now she’s a spitting image of her mom.” So, she looks like herself now. You’re going to make her look like her family members when you’re done.”
- “Sometimes they’ll have speech improvement just because their teeth come together in a proper position. Those sounds that are what we call dentoalveolar sounds or dentolabial sounds that rely on the position of your tongue to your teeth and your lips to your teeth, those will improve.”
- “I would say the advantage is then [THE PATIENT] would have a somewhat improved appearance at a younger age. It’s certainly not going to correct some of the major problems, but it may give ……. some confidence and help [THE PATIENT] feel a little bit better about …… for the next few years until [WE CAN] to do something more aggressive.”

#### Theme 8. Multiple Surgeries Are Needed

- “If you try to do too much surgery on that nose, you put in cartilage grafts, you try to really be aggressive in getting it perfect, then it will look good until you do your foundation …… [JAW] surgery and then it will look bad because you’ve changed the >foundation and then you’ve basically caused scarring and a whole bunch of other issues and you’ve potentially burned bridges because you weren’t thinking ahead of the other potential interventions that have to be done.”
- “I mean we try to combine things, minimize anesthesia as much as possible with these children who have multiple procedures.”

#### Theme 9. Orthodontics/Alveolar Bone Grafting And Orthognathic Surgery Affect Revision Surgical Planning

- “That’s where it comes down to talking with the family and the patient. [THE PATIENT IS] almost [AGE RANGE 9-11 YEARS], so I would definitely get a dental [INTRAORAL] image to see what—Some kids are missing the canine that’s there and that should be erupting through the cleft, so timing isn’t so much an issue. If [THE PATIENT] does have that tooth there and it’s ready, I would prefer to do the bone graft first, then wait a minimum of six months and then offer a lip-nose revision. If [THE PATIENT IS] really self-conscious and struggling in school and being teased, then I’d be more inclined to go ahead and offer a lip-nose revision.”
- “Well, [THE PATIENT] could wait……. doesn’t lose anything by waiting on doing these revisions, but I wouldn’t do them at the same time as the orthognathic [SURGERY] because it’s swollen and there’s so much other movement going on at the same time with the bite that you need to just do that operation and then let everything heal and let the swelling go down for six months and then you can do everything afterwards. You can do the final work on the nose, you could do any final work you want on the lip, but there’s too much going on with swelling that happens around the mouth at the time of the orthognathic surgery that you can’t really combine these other things with that.”

#### Theme 10. Treatments Are Impacted By Surgeon And Patient Resources

- “I think a lot of people [SURGEONS] make decisions based on the resources they have available and their experience. I know we’ve talked in the past about laser scar treatment and that’s not something you can get everywhere……a lot of people probably wouldn’t suggest laser, not because they’re looking at the pictures [FROM THE SAFS] differently or have a different set of information. They just have a different set of resources …… so I think that’s probably the bigger decision-making factor for a lot of people.”

A variety of other less-discussed sub-themes emerge from surgeons’ responses some of which were discussed earlier in different contexts. Examples of these sub-themes include setting realistic expectations for surgical outcomes with the patient and family and emphasizing that postoperatively their cooperation is important to ensure the desired result. Related to the surgeons and the surgery is the recognition that some diagnoses and treatments are standard, however, surgeons may differ in the specific surgical revision procedures that they perform.

Asymmetry is a common issue that can be fixed with surgery, the effects of surgery on breathing must be a primary concern, and not every clinical situation requires surgery. Surgeons are trained to expect the unexpected as some issues are discovered intraoperatively. Lastly, ethnicity as a patient characteristic also affects surgeons’ decision-making.

### (2) Primary Lip Repair Surgery

#### Theme 1. Use Of Nasal-Alveolar Molding (NAM) Has Pros And Cons And There Are Alternatives

Discussion around nasal alveolar molding (NAM) which is a specific type of infant orthopedic (IO) technique used immediately prior to lip repair is by far the single most-discussed theme with an extent three times that of the second-most discussed theme (Theme 2: Working with the Family). Since NAM is a leading treatment strategy for infants born with CL/P, it is not surprising it emerged as the foremost theme. Generally, the primary lip and nasal repair are done at the same time. Surgeons described the NAM as a common strategy for shaping the nose and specifically molding/elongating the columella prior to the lip repair. This is especially the case for a baby with a bilateral CL/P where the cleft segments are separated and there is a protrusive premaxilla. In such a case, soft tissue work becomes more difficult and special considerations of using infant orthopedics to realign the premaxilla prior to lip repair is preferred. The device includes a palatal plate with attached nasal stent(s) or struts that exert a ‘push’ force from the palatal plate to mold the nasal cartilage. Surgeons expressed the view that NAM may limit the number of future operations for a patient. It is best that treatment with NAM starts during the first few weeks after birth when nasal cartilage is most amenable to molding, however, NAM is just one possible IO appliance and there are others used by surgeons. Two other examples mentioned are the DynaCleft and the Latham appliance. The DynaCleft nasal elevator adheres to the forehead with a hook around the dome of the nasal cartilage to elevate (exert a pull force) and mold the cartilage. Advantages to this appliance are that it is more versatile because it can be used in conjunction with other IO appliances and the nasal molding can be initiated earlier in life than for the NAM. The Latham appliance is a pin-retained plate that requires a brief general anesthesia to be placed in the mouth. Surgeons’ choice of appliance may be family dependent. The NAM requires many clinic visits for adjustment and depending on the parents’ resources and ease of accessibility to the clinic, it may not be the best choice for certain families. Because the Latham appliance is fixed to the palate and requires less visits, it may be a better choice when clinic accessibility is an issue. Also, there may be improvements in feeding infants with these appliances.

Surgeons recognize some risks and drawbacks to NAM. These include skin irritation in the infant, and for both the clinician and parents, there is a significant time investment and a steep learning curve when the appliance is used. For example, clinicians need to make frequent adjustments to the palatal plate and nasal stent(s) of the appliance and parents need to be compliant with NAM for a successful outcome. Feeding is a possible risk initially, especially at the time that the nasal stents are placed but it is only a temporary risk until the parents become adept using the appliance.

- “I use nasoalveolar molding. I recommend it in cases where the lateral segments or particularly, the alveolar ridges aren’t lined up well, to help to approximate them, and to help improve the nasal shape before the time of surgery. And also, to help with feeding. I use it really whenever I feel that there’s a family that will be compliant with it because I feel like anyone that has a severe enough deformity, that the molding changes are warranted, as long as [THE] family can be compliant because it is an investment in time and money ……”

#### Theme 2. Working Closely With The Family Is Important

Working with the parents toward realistic expectations—expectations with regard to the surgery itself and the outcome of surgery is the second-most discussed theme. When discussing surgery, the conversation is kept non-technical and focuses on goals (e.g., making the lip continuous, improving the shape of the nasal tip), and what to expect after the surgery (e.g., expect a baby with a very different face). It is important to take steps to reassure the family regarding the immediate surgery—such as what happens when they arrive on the day of the surgery, where they will stay during the surgery, the duration of the surgery—and provide general support. There are parental support groups, for example, the American Cleft Palate Association’s CleftLine provides valued resources for questions regarding expectations around the surgery. Accommodating the family and letting them make decisions where possible (e.g., whether to use the NAM) is preferred. Overall, surgeons find that families generally view the outcome of lip repair surgery favorably.

- “They will feel like his nose is flatter even with the columella lengthening and they will feel like [THE PATIENT’S] upper lip is tight. But then they’ll love everything else, you know, [THE PATIENT WILL] look symmetric……look like his [HIS SIBLING] now and all that stuff.”

#### Theme 3. Diversity Among Patients Impacts Diagnosis, Planning, And Outcomes

Every infant requiring a lip repair presents a unique challenge. One learns from previous cases, but every case provides new challenges that may require a variation on the surgical technique. Surgeons find these surgical challenges inspiring. At times, only when the patient is asleep or anesthetized can a proper evaluation be accomplished. Age is a consideration in the infant—it is best for the infant to be strong, healthy, and gaining weight, and delaying surgery until these factors are achieved is important. Race is another factor that affects the surgical selection during the planning stage.

- “There are some benchmarks that we’ve been using……which is that [A] baby should weigh 10 pounds and have a hemoglobin of 10 and be at least 10 weeks old. I don’t know if there’s good hard data supporting that …… but generally speaking we’d like to get out of the perinatal period so [A] baby’s a little strong, demonstrate that they’re gaining weight, and that they’re healthy.”

#### Theme 4. Soft Tissue Work has Special Considerations

There are multidimensional aspects and nuances to the soft tissue work for an adequate lip repair when bone is involved. For example, in the patient with a bilateral CL/P who has a protrusive pre-maxilla repair may be technically difficult because stretching of tissues is involved to achieve an adequate repair. When repairing a wide cleft where a lot of bone is missing one surgeon described the surgical approach as “trampolining that soft tissue across this big gap” where eventually over time because of a lack of bone support the soft tissue sags.

Invariably, these infants will need a lip revision later in life but there is an awareness that further surgery increases the possibility of increased scarring leading to possible abnormal lip lengths and asymmetry.

- “So in … kids that I’m closing that have really wide gaps, a big hole, I don’t worry so much about giving them the perfect lip scar and I don’t worry so much about lengthening their columella cause I know I’m coming back and so I’d rather just get things together and let them heal and let the tissue stretch in general and then when I go and do the inevitable second operation [e.g., revision], that’s when I’ll do the fine tuning.”

#### Theme 5. Lip Repair Surgery Has Risks And Limitations

As expected, primary lip repair surgery is not entirely predictable and involves risks and the possibility of relapse. Structural changes occur as the infant ages and scarring becomes more obvious. Surgeons note the importance of properly placed incisions at the time of the lip repair to mitigate visible scarring later in life. When it comes to relapse, the nasal cartilage is particularly prone.

- “On a unilateral, I’d do a reasonably aggressive primary tip rhinoplasty. We try to realign the nostril shape or the alar. You can usually set the alar bases well and try to get the shape of the nostril well, but sometimes that septal portion, even if you get it sitting where you want it and you put some sutures in to hold it, I often think those sutures are there just to make me feel better about what it looks like at that moment and then a couple of weeks later, you’re like, “That didn’t really hold very well.” I don’t know that there’s a great way to make it stay because it already has a lot of memory to it as cartilage at that time point.”

#### Theme 6. Different Surgeons Take Different Approaches To Treatment

The surgeons differ on the technique they use for lip repair and that is partly dependent on their training. The techniques mentioned include the Fisher and Millard as well as modifications thereof.

- “I think it has a lot to do with your training and what technique you were exposed to during your training. The Fisher technique is just a ton of different points that you have to draw out and different flaps and actually a different technique to closing a cleft lip than the Millard …… but some people who know how to do both techniques base which technique they’re going to do on the anatomy of the cleft.”

#### Theme 7. Other

Other sub-themes mentioned for primary lip repair are the importance of ‘live infant examinations’ especially as it pertains to tactile information—manipulating the circumoral tissues is very informative. Telehealth was mentioned as having its advantages but being limited compared with ‘face to face’ examinations. While initial examinations can be conducted with telehealth, the need to examine an infant in person is imperative.

- “[COMMENT BY A SURGEON ON TELEHEALTH] ……. during the lockdown. I would say that the 2D photos were really the mainstay of that, and then to the extent we could use video to do movement, that was also very helpful. While that is useful for me in terms of surgical planning, I found that things like feeding discussions and discussions of how to place—I use this thing called DynaCleft that molds the nose, how to place that—were really limited by telehealth, and I often ended up bringing kids in at a later date just to go over things that really were hard to address by telehealth… I think those are things where communication back and forth between the family and me, the surgeon, is really important ……”

### (3) Agreement Among Surgeons’ Treatment Planning Decisions

#### Lip Revision

To assess surgeon agreement on diagnosis and treatment planning decisions, two approaches are used. First, recall that each of the eight surgeons (A to H in Table 5) completed the SAFS for one non-revision patient and they were not aware of which patients were non-revision. The surgeons’ recommendations for the non-revision patients were compared and they demonstrated substantial agreement in their recommendations. Specifically, seven of the eight doctors recommended no surgery though in some cases there was a recommendation for minor surgical work. Second, four surgeons (A to D) looked at the SAFS for one revision patient (Patient 1) and the other four (E to H) looked at the SAFS for a second revision patient (Patient 2). We examined whether the surgeons agreed on the diagnosis and treatment for the patient’s SAFS that they viewed. Each of the surgeons who reviewed their respective patient’s SAFS provided largely the same basic diagnoses/treatments. Table 5 lists the statements for each patient. An “x” indicates that the surgeon identified in the column header made the given statement. Because interviews were semi-structured with interviewer prompts varying by surgeon, the absence of an “x” does not indicate disagreement with a statement but rather absence of information. For Patient 1, all four surgeons recommended further lip revision and three recommended nasal tip surgery. One surgeon [D] felt the need for some immediate work on nasal asymmetry. For Patient 2, three of the four surgeons recommended a lip revision, and even the surgeon that did not [H] recommended nasal tip work which (s)he felt would further improve the patient’s appearance. Two surgeons mentioned the need to address nasal asymmetry which was not addressed by one surgeon [F].

**Table 5.** Surgeons’ agreement for lip revision diagnosis and treatment planning using the SAFS.

| **Diagnosis and Treatment Planning** |  |  |  |  |
| --- | --- | --- | --- | --- |
| **Statements by Surgeons (Patient 1)** | **Surgeon Codes** |  |  |  |
|  | A | B | C | D |
| No obvious scarring needing surgery | x |  |  |  |
| Lip height good | x |  |  |  |
| Dent in red lip so soft tissue deficient and lip revision recommended now | x | x | x | x |
| Upper lip too tight, causes pouting effect | x |  |  |  |
| Upper jaw set back, needs jaw surgery, orthognathic correction | x | x | x |  |
| Braces may be alternative to jaw surgery |  |  | x |  |
| Cheek puff/pucker does not warrant surgery | x |  |  |  |
| Nasal asymmetry, to be addressed later to correct nasal tip |  | x | x | x |
| Would do some nasal work now |  |  |  | x |
| **Statements by Surgeons (Patient 2)** | **Surgeon Codes** |  |  |  |
|  | E | F | G | H |
| Abnormal wet mucosa in upper lip should be inside mouth; upper lip abnormally thick | x |  | x |  |
| Skin tethering scar affecting upper lip, muscle surgery may be required | x |  |  |  |
| Prominent lip scar can be dealt with using fat graft |  | x |  |  |
| Nasal asymmetry between nostrils, bone graft and rhinoplasty needed | x |  | x | x |
| Septorhinoplasty needed when growth is complete | x |  |  | x |
| Lip revision not needed |  |  |  | x |

#### Lip Repair

Refusing to perform a lip repair in an infant is not an option, thus, the focus is on agreement around nuances of the repair. Four surgeons (A to D) evaluated the SAFS for one infant (Infant 1) and the other four (E to H) evaluated the SAFS for a second infant (Infant 2). Both infants had an unrepaired bilateral CL/P. The surgeons provided largely the same basic diagnoses/treatments for each infant (Table 6). For Infant 1, three of the four surgeons recommended lengthening the columella, removal of lower lip pits, and pre-surgical infant orthopedics prior to the surgery. Two surgeons also recommended some form of rhinoplasty to address asymmetry and nasal tip issues. For Infant 2, all four surgeons recommended rhinoplasty to address narrowing of alar bases, reshaping of nostrils, and lengthening of the columella and one surgeon recommended removal of an excess skin nubbin. No surgeon made a recommendation contrary to that of another.

**Table 6.** Surgeons’ agreement for lip repair diagnosis and treatment planning using the SAFS.

| **Diagnosis and Treatment Planning** |  |  |  |  |
| --- | --- | --- | --- | --- |
| **Statements by Surgeons (Patient 1)** | **Surgeon Codes** |  |  |  |
|  | A | B | C | D |
| Bilateral cleft lip repair needed | x |  | x | x |
| Bilateral cleft palate repair needed | x |  | x | x |
| Rhinoplasty (asymmetry, nasal tip) needed | x |  |  | x |
| Diminutive columella needs work | x |  | x | x |
| Protrusive premaxilla and prolabium needs work | x |  | x | x |
| Lower lip pits need removal | x |  | x | x |
| Pre-surgical orthopedic device needed (NAM or Latham) | x |  | x | x |
| **Statements by Surgeons (Patients 2)** | **Surgeon Codes** |  |  |  |
|  | E | F | G | H |
| Bilateral cleft lip repair needed | x | x | x | x |
| Bilateral cleft palate repair needed |  | x | x | x |
| Rhinoplasty indicated (alar bases should be narrower, reshape nostrils) | x | x | x | x |
| Excess skin nubbin | x |  |  |  |
| Columella/upper lip needs lengthening | x | x | x | x |

\* No comments for B

### (4) Experience Levels of Surgeons

The effect of the surgeons’ level of experience treating patients with CL/P was assessed using the second approach similar to the agreement assessment whereby four surgeons looked at the SAFS for revision Patient 1 and repair Infant 1, respectively, and the other four looked at the SAFS for revision Patient 2 and repair Infant 2. Four surgeons had five or more years of experience and four had less than five years of experience at the start of the study. Tables 7 and 8 list their quotations related to diagnosis and treatment planning for the revision and repair patients. For the revision Patient 1, both the experienced and less experienced surgeons largely agreed on diagnosis and treatment. The clearest difference lay in the less experienced surgeons recommending work on lip scarring whereas none of three more experienced surgeons made this recommendation. Both experienced and less experienced surgeons foresaw the need for jaw surgery, though the experienced surgeons provided more details on this. For the revision Patient 2, both subsets of surgeons largely agreed on the diagnoses, however, the less experienced surgeons recommended more aggressive surgeries. For the repair Infants 1, both experienced and less experienced surgeons again largely agreed on the diagnosis. There was agreement on the need for pre-surgical infant orthopedics, on the surgery, and on the need to excise lip pits; however, the experienced surgeons also recommended osteotomy and open tip rhinoplasty. For Infant 2, the surgeons all agreed on the diagnosis, and all but one, on the treatment—one of the experienced surgeons differed on whether to address the premaxilla and whether to recommend NAM.

**Table 7.** Assessment of surgeon experience for lip revision diagnosis and treatment planning using the SAFS.

| **Diagnostic Comments for Lip Revision Patient 1** |  |
| --- | --- |
| Experienced Surgeons | Less Experienced Surgeons |
| Cleft lip | Cleft lip |
| Previous lip repair | Previous lip repair |
| No previous revisions | Not discussed |
| Previous bone grafting | Not discussed |
| Previous cleft palate repair | Not discussed |
| Dent in red lip due to deficient soft tissue | Depression at the top of her lip, filler needed |
| Upper lip too tight, bottom lip loose | Need to slightly lengthen upper lip |
| Upper jaw set back, maxillary hypoplasia | Not discussed |
| Lower jaw needs to be set back | Not discussed |
| Maxillary advancement needed | Not discussed |
| Jaw (orthognathic) surgery likely needed | Orthognathic surgery needed |
| Strong forehead and chin with flat mid-face | Deficiency of mid-vault |
| Nasal asymmetry, asymmetrical alar cartilage | Nasal asymmetry, asymmetry of alar base |
| Septal deviation, tip asymmetry | Septal deviation, tip asymmetry |
| Scarring is good | Lip scarring needs work |
| Rhinoplasty needed later | Needs septorhinoplasty later |
| Lip muscle may have dehiscence some on right | Not discussed |
| Not discussed | A little bulging of the orbicularis muscle |
| **Diagnostic Comments for Lip Revision Patient 2** |  |
| Cleft lip | Bilateral cleft lip and cleft palate |
| Asymmetry of alar base | Nasal asymmetry between nostrils |
| Scar across lip but lip shape is good | Had previous lip revision but resecting runs risk of worse scarring. |
| No reason to address movement of lower lip | Upper lip movement good |
| Fat graft would improve lip | Botox possible |
| Opposes aggressive lip surgery | Favors lip revision operation |
|  | Wet mucosa outside rather than inside mouth needs removal. |
|  | Upper lip abnormally thick |
|  | Bone graft recommended for nose |
|  | Final septorhinoplasty when growth done |

**Table 8.** Assessment of surgeon experience for lip repair diagnosis and treatment planning using the SAFS.

| **Diagnostic Comments for Lip Repair Patient 1** |  |
| --- | --- |
| Experienced surgeons | Less experienced surgeons |
| Bilateral complete cleft lip and palate | Complete bilateral cleft lip |
| Presurgical orthopedics (NAM or Latham) strongly recommended to deal with fly-away premaxilla | Prominent premaxilla, orthopedics recommended (NAM or Latham) |
| Displaced prolabium requires surgery to narrow the prolabial flap and bring back together the upper lip muscle and skin, repairing mucosa and gums | Surgery needed for bilateral cleft lip (less detailed discussion) |
| This involves an osteotomy, repositioning maxilla not to jut so far forward | Osteotomy or jaw surgery not mentioned |
| Bilateral open-tip rhinoplasty involving dissecting out cartilage in nose and repositioning it | Some nasal asymmetry noted but rhinoplasty not mentioned |
| Very short columella | Very short columella |
| Lip pits need to be surgically excised | Lower lip pits need excising |
| **Diagnostic comments for Lip Repair Patient 2** |  |
| Straightforward bilateral cleft lip repair | Complete bilateral cleft lip and palate |
| One experienced doctor views premaxilla favorable, jaw not sticking out, but other experienced doctor say patient has locked-out premaxilla. | Premaxilla not bad. |
| One experienced doctor says presurgical orthopedics not needed, but other experienced doctor recommends NAM | Favorable to NAM but dialog is unclear whether doctor would recommend for this patient. |
| Skin nubbin sticking out but no bone | Skin nubbin not addressed specifically but may be part of cleft repair. |
| Nose, alar bases should be a little narrower | Good nasal symmetry but alar bases are wide, need narrowing; cleft rhinoplasty recommended |
| Columella a little short, needs lengthening | Columella a little short, needs lengthening |

**Table 9.** Lip revision checklist.

| Lip Revision Checklist |  |
| --- | --- |
| Question | Answers/Considerations |
| **From your perspective, is this the best time for**<br>Lip revision<br>Nasal revision<br>Alveolar bone grafting<br>Jaw surgery<br>* Might previous lip and nasal surgery impact your outcome?<br>* Could speech be affected by the planned surgery? |  |
| **Surgical risks.**<br>Is your patient particularly prone to:<br>Excess bleeding, infection, occurrence of necrotic tissue<br>Worsening of scars<br>Development of hypertrophic tissue<br>Increased hypernasality<br>The need for jaw surgery (midfacial advancement)<br>Relapse<br>If you said yes to any of these, what leads you to believe this could be a specific risk for your patient?<br>How aggressive are your surgical plans? (Rate from 1-10, with 10 being most aggressive)<br>Why does your plan deserve this rating? |  |
| **Patient and family goals.**<br>What are the patient's concerns/goals (elicit in own words)?<br>What are the family's concerns/goals (elicit in own words)?<br>Do the patient and family concerns/goals match? If not how will you reconcile differences?<br>What areas of the face bother the patient?<br>Are there any psychological concerns from the patient and/or family perspective? How will you manage these concerns? |  |
| **Which specific facial areas do you plan to address/improve with** |  |
| **surgery and describe why?**<br>Upper lip length? Describe.<br>Upper lip width? Describe.<br>Upper lip symmetry? Describe.<br>Increase/decrease the upper lip thickness? Describe.<br>Lengthen the columella? Describe.<br>Restore cupid's bow? Describe.<br>Adjust lower lip form? Describe.<br>Any additional areas? |  |
| **Which specific facial movements do you plan to address/improve with surgery and why?**<br>Upper lip vertical movement? Describe.<br>Upper lip horizontal movement? Describe.<br>Lower lip movement? Describe.<br>Any other facial movements? |  |
| **Plans for the actual surgery (pre-surgical planning).**<br>Do you have adequate muscle for the revision? Describe.<br>Map out the surgical maneuvers for:<br>Soft tissue lip/augmentation<br>Soft tissue nose/augmentation<br>Manipulating/grafting nasal cartilage<br>Alveolar bone grafting, if it is to be done at the same time |  |
| **Scar tissue.**<br>Do you plan to revise the scar at the time of the surgery? How will you manage the effect of:<br>Prior scarring and its effect on the outcome?<br>Multiple previous surgeries and its effect on the muscle and the outcome?<br>Multiple previous surgeries and its effect on the muscle?<br>Other non-surgical options? e.g., laser, fat injection, Botox.<br>How will the patient's age impact your surgery?<br>Will you make direct measurements/markings on the patient's face while the patient is under anesthesia?<br>Will you reconfirm the surgical plan with the family on the day of, and immediately before, the surgery? |  |
| **Do you think that additional surgeries will be needed for your** |  |
| **patient in the future? If yes, expand.**<br>**Could future orthognathic surgery needs affect the revision outcomes? If so, describe.** |  |
| **Are there treatments unavailable to you in your clinic e.g., laser therapy, that might be beneficial alternative for this patient's soft tissue problems?** |  |
| **Have you set realistic expectations for the patient/family? Please describe how you set those expectations?** |  |
| **How have you assessed the family's comprehension and motivation to comply with post-surgical care at home?** |  |
| **Have you arranged for additional support to assist the family in executing a post-surgical home care plan?** |  |
| **How is the patient's ethnicity affecting your surgical plans?** |  |

## DISCUSSION

A wide variety of common themes emerged from the interviews with surgeons regarding the decision-making process for CL/P surgery that reflect the pervasiveness of these decisions. At nearly every stage of treatment decisions are made: Some in conjunction with patients/families, others with the CL/P team, and still others by the surgeon alone. Reviewing the numerous and varied considerations that characterize the decision process is useful both for trainees and experienced practitioners. For surgeons in training, a methodical and inclusive review of the considerations is of obvious benefit. And for surgeons actively involved in cleft care, a comprehensive list of considerations that comprise each surgical decision is helpful—if for no other reason than ensuring each decision is well-reasoned and given thorough weight. To this end, the surgical considerations that emerged provide useful and comprehensive information to populate checklists (Tables 10 &11) for the CL/P surgeon and her team that would be a valuable resource and a first step for those surgeons seeking to develop a structured process for making surgical decisions. A comparative review of the surgeons’ ‘top five’ themes demonstrates a difference in emphasis of the considerations by patient type (revision and repair); however, three themes emerged as common to both types, ‘working closely with the parents/family’, ‘risks and limitations of surgery’, and ‘soft tissue considerations/muscle repair’. Soft tissue and other considerations were explored in depth earlier, however, interactions with the patient/family and the risks and limitations of surgery warrant further consideration.

**Table 10.** Lip repair checklist.

| Lip Repair Checklist |  |
| --- | --- |
| Question | Answers/Considerations |
| **Pre-surgical infant orthopedics (IO).**<br>Do you plan to use an IO appliance. If yes, which will you use and why?<br>NAM<br>Latham<br>DynaCleft<br>Another type of IO appliance (please specify).<br>Will you combine appliances? Why?<br>Did you discuss with the family the advantages and disadvantages of the chosen appliance(s)?<br>Did you or your team conduct an assessment to discern whether the family can comply with your recommended IO appliance? Or would another appliance would be more suitable for their home situation?<br>What ideal age in months will you start IO on this infant?<br>What are your anatomical expectations for stopping IO treatment on this infant?<br>Do you plan to use the IO appliance after the surgery?<br>Did you provide verbal and written instructions on using the appliance to the family? How have you confirmed that the family understands the instructions? (e.g., teach-back method? Other?) |  |
| **Realistic expectations for the family.**<br>How have you set realistic expectations for the family?<br>What are your goals for the lip repair surgery?<br>Have you described the goals to the family in non-technical terms?<br>Have you described to the family how their infant will look after the surgery?<br>Have you reassured the family regarding the surgery itself, e.g., what to expect on the day, where they will stay, who will update them |  |
| on progress of the surgery, etc?<br>Have you provided the family with information on contact information for support groups, e.g., ACPA's CleftLine? |  |
| **Surgery and surgical risks.**<br>Is the infant healthy enough to have the surgery? What benchmarks have you used to determine the infant's overall health?<br>What technique for lip repair will you use and why?<br>How do you plan to minimize the effects of scarring of the lip?<br>Is the patient likely to have greater than expected scarring after surgery and healing? And why do you expect this?<br>Did you make a preliminary assessment as to whether revision surgery will be needed after the initial lip repair? And if so, how did you make that determination?<br>Do you expect relapse of the nasal septal cartilage position as the infant ages? If so, why?<br>When you plan the technique for lip repair surgery, do you consider how it could affect lip movement? |  |

Decisions made by surgeons in conjunction with the patient and parent/caregiver—shared decision making—occurs when patients/parents and providers collaborate to develop a mutually agreed treatment plan.^24^ It brings quality to the decision-making process over and above the decision itself.^25^ In a systematic literature review of shared decision making on patient choice for elective general surgical procedures, Boss and co-workers (2016)^26^ found that although this shared process may reduce or have no impact on patient choice for elective surgery, it may promote a more positive health care experience and decision-making process for the patients. Lip revision in patients with CL/P is an elective surgery, and these patients and their families want to participate in surgical decisions but have limited understanding of their facial difference and the surgical indications.^27^ Surgeons must educate their patients and facilitate the decision-making process.^27^ The surgeons in this study emphasized the importance of doing just that.

When considering the ‘risk and limitations of surgery’, once again the focus is different depending on patient type. Specifically, primary lip repair in an infant is a forgone conclusion and surgeons can only hope to lessen and/or avoid risks when possible. Alternatively, the choice of doing a lip revision is intertwined with the surgical consequences in the form of the risks and the limitations. In this instance, the patient/family and surgeon have a choice of *whether* and *when* to perform the surgery based on the surgeon’s expert decisions. Cooper et al (2020)^28^ in a sample of 882 patients found that 15% of patients had deficits in knowledge of their diagnosis and /or procedure. Once surgeons identify the surgical risks, knowledge deficits in patients/families surrounding the risks and limitations of the surgery must be avoided and identifying patients that are particularly prone to such deficits is important to facilitate high-quality decisions by patients/families and surgeons alike. For surgeons, adequate surgical preparation and planning can help avoid negative surprises associated with the surgical procedure. A thorough clinical examination of the patient coupled with a systematic planning process like the SAFS and checklists of factors to be addressed as demonstrated in this study help to maximize positive outcomes. These checklists can be tailored for use by any clinician.

Although not a main aim of the study, an interesting finding was that the surgeons were consistent in their recommendations. Past research has demonstrated that when surgeons view the same set of patients, in general they disagree in their recommendation for lip revision [11].^11^ In this study, surgeons agreed on most of the clinical observational findings for the same child that they viewed. Perhaps this type of in-depth analysis based on a systematic assessment of extensive images by surgeons improves agreement at the level of clinical diagnostic observations. However, the final recommendation for ‘choosing to do a revision’ may lead to disagreement among surgeons because many other factors are at play that include input from the patient and family regarding the desire to have a surgery; suspicion that scarring may worsen because of multiple surgeries; the degree of surgical experience; confidence that one can deliver a successful result; and other variables. These factors that are unrelated to the surgeon’s actual diagnostic findings may have a greater bearing on the discord among surgeons when the final recommendation for revision surgery is made. Finally, although there were some minor differences in diagnostic findings between the experienced and less experienced surgeons, surgical experience was not a big factor in the surgical decisions chosen for the patients.

### Other Considerations

In March 2020, with the advent of the COVID-19 pandemic Tufts University and University of North Carolina at Chapel Hill suspended research activities for several months. During this time, only virtual, non-patient contact, research activities were approved. At that time, patient recruitment and patient data-collection for the SAFS were complete. The SAFS presentations to the surgeons, which prior to the pandemic were conducted in-person at the surgeons’ respective Craniofacial Centers, were switched to virtual presentations. This change allowed an evaluation of a virtual ‘remote’ platform for treatment planning. Thirty virtual SAFS presentations were conducted, and every surgeon participated in at least one. Most of the surgeons viewed the presentations on a laptop, some used a desktop, and one surgeon viewed one presentation on an iPhone. They rated their experience along a Likert scale as follows: 1 = “unsatisfactory”, 2 = “satisfactory”, 3 = “very satisfactory, I prefer this method”. Overall, the surgeons were very positive on the virtual presentations. They rated 28 of the 30 virtual presentations as a ‘3’ and two were rated a ‘2’. Positive comments were that the process for the presentations felt comfortable; more convenient especially for scheduling viewings; just as efficient as the in-person presentations with the ability to conduct presentations anywhere and anytime; and cost-effective—no travel time needed. Negative comments were the lack of intraoral images and inability to control and move the 3D images; however, for the latter the viewer can instruct the presenter to do this. Also, for three presentations there were minor technical problems that were easily fixed.

There were a few caveats in this study. The surgeon participants were from four large health centers on the east coast and there may be a concern of external validity; however, the surgeons can be said to represent a good balance based on their demographic factors and surgical experience. All were credentialed medical professionals, and all decisions and interviews were based on the SAFS method. Finally, the themes were ranked by the number of associated quotations, and we chose to infer importance based on this rank order. We considered this a logical inference; however, the surgical checklists complied from this research include all considerations without inferring importance which, in this instance, is left to the surgeon.

## Data Availability

All data produced in the present work are contained in the manuscript.

