## Supplemental File 1 for "Decision Considerations and Strategies for Lip Surgery in Patients with Cleft lip/Palate: A Qualitative Study"

### Targeting Surgeons' Decision-Making for Cleft Lip Surgery

**NIDCR Protocol Number: 15-069-E**

**NIDCR Grant Number: U01 DE024503**

**Principal Investigator: Carroll Ann Trotman, BDS, MA, MS**

**NIDCR Program Official: Melissa Riddle, PhD**

**NIDCR Medical Monitor: Kevin McBryde, MD**

**Draft or Version Number: 3.0**

**10 Jan 2018**

#### STATEMENT OF COMPLIANCE

The study will be conducted in accordance with the International Conference on Harmonisation guidelines for Good Clinical Practice (ICH E6), the Code of Federal Regulations on the Protection of Human Subjects (45 CFR Part 46), and the NIDCR Clinical Terms of Award. All personnel involved in the conduct of this study have completed human subjects protection training.

#### SIGNATURE PAGE

The signature below constitutes the approval of this protocol and the attachments, and provides the necessary assurances that this study will be conducted according to all stipulations of the protocol, including all statements regarding confidentiality, and according to local legal and regulatory requirements and applicable US federal regulations and ICH guidelines.

Principal Investigator or Clinical Site Investigator:

Signed: [signature] Date: 01/19/2018

Name: Carroll Ann Trotman, BDS, MA, MS

Title: Principal Investigator

#### LIST OF ABBREVIATIONS AND SPECIAL TERMS

| | |
|---|---|
| AE | Adverse Event/Adverse Experience |
| Caregiver | Individual responsible for caring for a minor patient. This term includes parents as well as other individuals who play that role. |
| CFR | Code of Federal Regulations |
| CRF | Case Report Form |
| CRO | Contract Research Organization |
| DCC | Data Coordinating Center |
| DHHS | Department of Health and Human Services |
| DSMB | Data and Safety Monitoring Board |
| eCRF | Electronic Case Report Form |
| FALS | Facial Animation Lab Staff |
| FWA | Federal-wide Assurance |
| GCP | Good Clinical Practice |
| Guardian | Legal signatory for minor patient. This term includes parents as well as other individuals with custodial and legal responsibilities. |
| HIPAA | Health Insurance Portability and Accountability Act |
| ICF | Informed Consent Form |
| ICH | International Conference on Harmonisation |
| ICMJE | International Committee of Medical Journal Editors |
| IDI | In-Depth Interview |
| IRB | Institutional Review Board |
| MedDRA® | Medical Dictionary for Regulatory Activities |
| MOP | Manual of Procedures |
| N | Number (typically refers to subjects) |
| NIDCR | National Institute of Dental and Craniofacial Research, NIH, DHHS |
| NIH | National Institutes of Health |
| OCTOM | Office of Clinical Trials Operations and Management, NIDCR, NIH |
| OHRP | Office for Human Research Protections |
| Patients | Individuals providing the photography and video data to be used by the surgeon-raters. In this study the patients are not the subjects of the SAFS Intervention, nor are they directly affected by the intervention. |
| PHI | Protected Health Information |
| PI | Principal Investigator |

| | |
|---|---|
| QA | Quality Assurance |
| QC | Quality Control |
| SAE | Serious Adverse Event/Serious Adverse Experience |
| SAFS | Systematic Assessment for Facial Surgery. (The surgeon-raters observe still photographs, videos, and quantitative information of patients engaging in various facial activities. During the review, the surgeon-raters respond to a series of pre-specified questions about the images.) |
| Surgeon-raters | Surgeons receiving the study intervention (SAFS). Surgeon-raters are the subjects in the study. It is their behavior that is predicted to be affected by engaging in the SAFS intervention. |
| UP | Unanticipated Problem |
| US | United States |
| WHO | World Health Organization |

#### PROTOCOL SUMMARY

**Title:** *Targeting Surgeons' Decision-Making for Lip Surgery*

**Précis:** This study includes: (1) A group of surgeon-raters who will review images and quantitative information of patients with Cleft Lip and Palate (CL/P); and (2) Two prospective groups/cohorts of patients with cleft lip/palate (CL/P) – one cohort recommended to have secondary lip revision surgery and another slated to have primary lip repair surgery.

The study will assign surgeon-raters to evaluate patients who are scheduled to be treated at another participating study site. Each surgeon-rater will be assigned to hypothetically assess several patients from both cohorts for surgery. Surgeon-raters will use a novel pre-surgical planning system for this evaluation – the Systematic Assessment for Facial Surgery (SAFS) Intervention. Subsequent to each SAFS, surgeon-raters will participate in an In-depth interview (IDI), designed to illuminate the surgeon-raters' decision making process. This study will determine how the SAFS Intervention affects surgeons' decision-making and treatment plans for surgery.

**Objectives:** The primary objectives of this study are:

1. To qualitatively assess how surgeon-raters integrate the SAFS's objective measures and visual aids with the SAFS's systematic subjective assessment in the decision-making process for the clinical surgical procedures of lip revision.

2. To qualitatively assess how surgeon-raters integrate the SAFS's objective measures and visual aids with the SAFS's systematic subjective assessment in the decision-making process for the clinical surgical procedures of primary lip repair.

The secondary objectives of this study are:

3. To quantitatively assess the extent to which the SAFS changes surgeon-raters' problem list and treatment planning goals for lip revision.

4. To quantitatively assess the extent to which the SAFS changes surgeon-raters' problem list and treatment planning goals for primary lip repair.

5. To quantitatively assess the extent to which the SAFS changes surgeon-raters' problem list and treatment planning goals for lip revision as a function of surgical expertise.

| | |
|---|---|
| **Population(s):** | (1) A group of surgeons (n=up to 10) who may perform lip revision and/or lip repair surgery on their patients with CL/P (outside of the study) and who, as part of the study, will perform a structured evaluation of assigned patients' photographic, video, and quantitative assessments.<br>(2) A group of patients (ages 4-21 years) with repaired CL/P who are recommended lip revision [lip revision group; n=32] and infants (ages birth – 8 months) born with a CL/P who need a primary lip repair [primary lip repair group; n=16] |
| **Number of Sites:** | 6 |
| **Description of SAFS Intervention:** | Each surgeon-rater is shown a prescribed series of still and video images of each of his/her assigned patients. These images are accompanied by quantitative measures including (1) 3D static facial image data compared with mean control static image data and (2) 3D dynamic and statistical modeling of patients' mean facial movements compared with mean control movement data. The surgeon-raters evaluate the materials by responding to a series of standardized questions, designed to quantify facial disability for the treatment planning of lip surgery and for assessing surgical outcomes. |
| **Study Duration:** | 24 months |
| **Participation Duration:** | Patients: up to 3 months; Surgeon-raters: up to 1 year 11 months |
| **Estimated Time to Complete Enrollment:** | 21 months |

#### Schematic of Study Design:

**Prior to Patient Enrollment**

| Patients presenting for surgical consult are screened for eligibility | Consent to review medical records is obtained |
|---|---|

↓

**Recruitment and Consent**

| *Up to10 surgeon-raters with lip revision and/or lip repair expertise* | 16 patients scheduled for primary lip repair surgery and 32 patients scheduled for lip revision surgery |
|---|---|

↓

**Visit 1 (3 months to 1 day prior) and Visit 2 (if needed up to 1 day prior)**

2D and 3D still and video images are obtained from each patient before surgery

↓

***Intervention/Decision Visit (this visit could extend up to 3 days)***

*Surgeon-rater completes baseline reviews of a patient's materials, completes the SAFS intervention, and then completes the IDI*

### 1 KEY ROLES AND CONTACT INFORMATION

**Principal Investigator:**

Dr. Carroll Ann Trotman, BDS, MA, MS
Professor, Chair & Program Director
Department of Orthodontics
Interim Chair, Advanced and Graduate Education
Tufts University School of Dental Medicine
1 Kneeland Street, Boston MA 02111

**Medical Monitor:**

Kevin McBryde, MD
Medical Officer
The National Institute of Dental and Craniofacial Research
6701 Democracy Blvd. RM: 638
Bethesda, MD 20892

**NIDCR Program Official:**

David B. Clark, DrPH
Behavioral and Social Sciences Research Branch
National Institute of Dental and Craniofacial Research
National Institutes of Health
6701 Democracy Blvd. RM:650
Bethesda, MD 20892-4878

**Clinical Site Investigators:**

Carroll Ann Trotman, BDS, MA, MS
Tufts University School of Dental Medicine
1 Kneeland Street
Boston, MA 02111
Office Phone Number: 617- 636-0846
Facial Animation Lab Phone Numbers:
617-636-0347; 617-636-3591
Fax Number: 617-636-2740


Richard Bruun, DDS
Boston Children's Hospital
300 Longwood Avenue
Boston, MA 02115
Phone Number: 617-355-3375
Fax Number : 617-730-0478


Eric Liao, MD, PhD
Massachusetts General Hospital
55 Fruit Street
Boston, MA 02114
Phone Number: 617-724-9922
Fax Number: 617-726-8089


Ceib Phillips, PhD, MPH
University of North Carolina at Chapel Hill
CB 7450 Brauer Hall
Chapel Hill, NC 27599
Office Phone Number: 919-537-3373

Facial Animation Lab Phone Number: 919-537-3207

Fax Number: 919-843-8864

| | |
|---|---|
| **Data Coordinating Center:** | Tufts University School of Dental Medicine |
| **Facial Animation Labs:** | Tufts University School of Dental Medicine<br>University of North Carolina at Chapel Hill |
| **Other Key Personnel:** | M. Elizabeth Bennett, PhD, Health Psychologist<br>Clarity Consulting<br>Julian Faraway, PhD, Statistician |

### 2 INTRODUCTION: BACKGROUND INFORMATION AND SCIENTIFIC RATIONALE

#### 2.1 Background Information

*Health Problem*

Cleft Lip/Palate (CL/P) is among the most common of all birth defects, occurring once in every 600 to 800 births. Surgeons perform an initial (primary) repair of the cleft lip and nose to correct it. Unfortunately, the impact of the primary surgeries on facial soft tissue function or movement and facial form is highly variable.[1-3] Many patients remain with significant impairment in facial movement[1] and disfigurement in facial form,[2] and require additional, sometimes multiple, revision surgeries to improve their esthetic results.[1,3] In fact, revision surgery after the primary lip repair is the rule rather than the exception. Consequently, the resultant burden of care is great: (1) Costs for treatment expenses, ancillary services, time lost at work, and indirect costs such as the health/emotional well-being of the child and caregivers are high, and have increased over time;[4] (2) Parents report that the quality of mother-infant interactions is adversely affected,[5] that their children develop psychological problems because of their facial appearance,[5] and that later in life, many wish to have additional surgery;[6] (3) Patients with facial stigmata such as CL/P experience anxieties and awkward moments during social interactions with non-stigmatized individuals;[7] and (4) Less successful surgeries may impact patients socioeconomically in the form of diminished income and educational accomplishments compared with their non-cleft counterparts.[8-9]

*Standardized Assessment for Facial Surgery (SAFS) Intervention*

Our research group has developed a unique set of quantitative dynamic and static measures and quantitative visual aids for the evaluation of facial soft tissue function and form.[1-3,10-19] These measures include (1) 3D dynamic and statistical modeling of patients' mean facial movements and (2) 3D static facial image data, both compared with mean control movement and static image data, respectively. Ultimately, we developed a systematic evaluation method utilizing the objective measures and surgeons' subjective assessments to quantify facial disability for the treatment planning of lip surgery and assessing surgical outcomes.

*Relevant Research*

Our completed research projects have involved seven surgeons experienced in the treatment of patients with CL/P who provided on-going feedback regarding the utility and refinement of the SAFS Intervention for use in infants, older children, and adolescents.[15] This research highlighted the need for objective measures. For example, when the surgeons subjectively (qualitatively) evaluated patients for lip revision surgery, there was very poor agreement as to whether or not the patients should have a lip revision (Kappa values ranged from 0 to 0.57). Furthermore, even when some patients did receive a revision, the surgeons disagreed as to the success of the surgery—many went on to recommend additional revision surgery.[15] The only other large scale trial on nasolabial esthetics in CL/P patients, the Eurocleft Intercenter Study,[20] also used subjective ratings of the nasolabial region, and similar to our findings,[21] these researchers noted a low level of agreement among examiners, and an even greater level of complexity and discordance when subjective ratings were used to assess moving images.[20,21] These difficulties underscore the conclusion of the Eurocleft Study that assessment of the nasolabial region is "a key area for further research" and supports the need for objective measures of facial disability in facial animated behaviors to supplement surgeon's subjective clinical evaluations.

#### 2.2 Rationale

The standard-of-care to evaluate patients for lip revision surgery relies on a subjective assessment by the surgeon of the static face. The important role of function or movement generally has been given far less consideration mainly because of the challenges faced by surgeons (e.g., the amount/quality of the tissue available to alter movement). Presently, even when surgeons do attempt to assess function, they do so in a subjective manner because there are no quantitative/visual aids to incorporate functional assessment into their treatment planning and decisions regarding lip surgery.

The SAFS approach proposed here has been refined sufficiently with surgeon feedback to allow surgeons to broaden their "vista" of the patients' 'movement and form' problems. Potentially, having identified a movement/form problem(s), surgeons could contemplate what needs to be done to improve patient specific problem(s). Importantly, pilot studies demonstrated that the SAFS had a definite impact on surgeons' decisions for lip revision: Surgeons substantially, but variably, changed their problem list and treatment planning goals.[19] Thus, given that revision surgery is very common after the primary lip repair, an additional goal of this study is to understand surgeons' decision-making with the use of the SAFS, to determine surgeons' goals and expectations for primary lip repair surgery, and to understand the surgical limitations that may lead to subsequent revision surgery.

#### 2.3 Potential Risks and Benefits

The potential risks and benefits are described below.

##### 2.3.1 *Potential Risks for Patient Subjects*

The potential known risks for patient subjects in this study include the following:

(1) Breach of confidentiality. All patient subjects will be identified by a unique ID number. The document linking a study subject's name and ID number will be maintained in a linkage file accessible only by research staff and stored in a locked cabinet in a locked room. All study files will be password protected with specific permission levels for access. Consent will be obtained from patient subjects for

their photos and videos to be used in presentations and/or publications. Only aggregate statistics will be presented.

(2) Fatigue from session length. Subjects will be given breaks to rest, snack, and use the restroom, etc., in order to minimize their discomfort.

(3) Allergic reaction to adhesive used to place markers. All subjects will be tested by placing a small amount of the adhesive on the finger, the back of the hand and/or the upper arm and asked to wait for 20 minutes to see if there is a reaction. If any subject does have a reaction, the research team will treat the affected area with 1% hydrocortisone cream. The team will then use medical tape to help the dots stick to the participant's face.

##### 2.3.2 *Potential Risks for Surgeon-Rater Subjects*

The potential known risks for surgeon-raters in this study include the following:

(1) Breach of confidentiality. All surgeon-rater subjects will be identified by a unique ID number. The document linking a surgeon-rater subject's name and ID number will be maintained in a linkage file accessible only by research staff and stored in a locked cabinet in a locked room. All study files will be password protected with specific permission levels for access. Only aggregate statistics will be presented.

##### 2.3.3 *Potential Benefits for Patient Subjects*

Participation in this study provides no direct benefit to the patient subjects. The information gained from this study will potentially enhance surgical planning for children with unilateral or bilateral complete non-syndromic cleft lip.

##### 2.3.4 *Potential Benefits for Surgeon-Rater Subjects*

The information gained from this study will potentially enhance surgical planning for children with unilateral or bilateral complete non-syndromic cleft lip.

### 3 OBJECTIVES

#### 3.1 Study Objectives

The primary objectives of this study are:

1. To qualitatively assess how surgeon-raters integrate the SAFS's objective measures and visual aids with the SAFS's systematic subjective assessment in the decision-making process for the clinical surgical procedures of lip revision.
2. To qualitatively assess how surgeon-raters integrate the SAFS's objective measures and visual aids with the SAFS's systematic subjective assessment in the decision-making process for the clinical surgical procedures of primary lip repair.

Secondary objectives are:

3. To quantitatively assess the extent to which the SAFS changes surgeon-raters' problem list and treatment planning goals for lip revision.
4. To quantitatively assess the extent to which the SAFS changes surgeon-raters' problem list and treatment planning goals for primary lip repair.
5. To quantitatively assess the extent to which the SAFS changes surgeon-raters' problem list and treatment planning goals for lip revision as a function of surgical expertise.

#### 3.2 Study Outcome Measures

##### 3.2.1 *Primary Outcome Measures*

The primary outcome measures of the study are:

1. Surgeon-raters' decision making in lip revision

   Outcomes are based on transcribing the In-Depth-Interviews (IDIs)[22] that will be conducted with the surgeon-raters; coding the transcript with an a priori structure in mind (using the interview format as a starting point) but then using Grounded Theory[23-37] to guide further coding in order to capitalize on themes that emerge that may not conform to the interview format. Themes will be developed (from the interview format and/or the emergent themes) that we will group by frequency as well as to their relevance to hypothesis generation as to how surgeon-raters might learn about and integrate patient data over time.

2. Surgeon-raters' decision making in primary lip repair

Outcomes are based on transcribing the IDIs[22] that will be conducted with the surgeon-raters; coding the transcript with an a priori structure in mind (using the interview format as a starting point) but then using Grounded Theory[23-37] to guide further coding in order to capitalize on themes that emerge that may not conform to the structured interview format. Themes will be developed (from the interview format and/or the emergent themes) that we will group by frequency as well as to their relevance to hypothesis generation as to how surgeon-raters might learn about and integrate patient data over time.

This thematic and frequency analysis will be conducted after the SAFS Intervention is conducted with the surgeons for those patients who are in need of lip revision surgery to address objective 1, as well as after the SAFS Intervention is conducted with the surgeons for those patients in need of primary lip repair surgery to address objective 2.

##### 3.2.2 *Secondary Outcome Measures*

The secondary outcomes of the study are:

1. The quantitative assessment of the extent to which the SAFS changes surgeon-raters' problem list and treatment planning goals for lip revision.

   Nominal outcome of the effect of the SAFS on surgeon-raters' treatment plans/goals in terms of whether the surgeon-rater changes his problem list and goals for lip revision.

2. The quantitative assessment of the extent to which the SAFS changes surgeons' problem list and treatment planning goals for primary lip repair.

   Nominal outcome of the effect of the SAFS on surgeon-raters' treatment plans/goals in terms of whether and how much the surgeon-rater changes his problem list and goals for primary lip repair.

3. The quantitative assessment of the extent to which the SAFS changes surgeon-raters' problem list and treatment planning goals for lip revision as a function of surgical expertise.

   Nominal outcome of the effect of the SAFS on surgeon-raters' treatment plans/goals in terms of whether the surgeon-rater changes his problem list and goals for lip revision based on the length of surgical experience (in years) of treating patients with cleft lip and palate.

### 4 STUDY DESIGN

This study will assess the impact of a surgical planning system on surgeon-raters' treatment decisions. The study will be conducted at six Craniofacial Centers serving as the clinical recruitment sites, and one Center serving as the data coordinating center (DCC). The recruitment centers are: The University of North Carolina (UNC), Wake Forest Baptist Health Craniofacial Center (WFB), Boston Children's Hospital (BCH), Massachusetts General Hospital (MGH), Shriners Hospitals for Children Boston (SHC-BOS) and Tufts Medical Center (TMC). Tufts University School of Dental Medicine (TUSDM) will serve as the data-processing and management center. Facial Animation Laboratories will be located at UNC and TUSDM.

At each Craniofacial Center, the operating surgeon will make the initial clinical decision to perform lip surgery on patients with CL/P. After the surgeons have made this decision, two patient groups will be enrolled: Individuals (age range=4 to 21 years) scheduled for lip revision surgery (n=32); and infants (age range=birth to 8 months) scheduled for primary lip repair (n=16).

For each lip revision and primary lip repair group, the patients will be recruited on or after the time they present for treatment and before surgery. Patients who are identified during screening will have their medical history data reviewed to determine study eligibility. Those qualified for the study will be scheduled for the data collection visit(s). Consent will be collected at a combined qualification and data collection visit.

Facial animation data (static 3D and 2D and 3D video imaging) will be collected from all patients on one to two separate pre-surgery visits. The pre-surgery visits will occur at no greater than 3 months before the scheduled surgery, and if needed, a second visit to complete data collection will occur up to 1 day before the surgery for both patient groups. The pre-surgery data will be used to compile the SAFS.

Facial animation data for patients from BCH, MGH, SHC-BOS and TMC will be obtained at the Facial Animation Laboratory at TUSDM. Likewise, the Facial Animation Laboratory at UNC will capture data for patients from WFB and UNC.

Surgeon-raters will be assigned to complete the SAFS on imaging and quantitative data for patients who are slated to be treated at another study site (i.e., patients who are not their own).

Specifically, each of up to ten surgeon-raters in the study will complete the SAFS for four patients slated to have a lip revision and two patients slated to have a primary lip repair.

The SAFS consists of data that will allow the surgeon-raters to conduct an evaluation utilizing objective measures of 3D facial soft tissue movements and a systematic subjective assessment to quantify facial disability for the treatment planning of the lip surgery.

All the participating surgeon-raters will be trained on and use the SAFS for the first time in this study; all have high volume surgical practices devoted to the care of patients with CL/P. They were selected based on their different levels of surgical experience so as to obtain a broad range of feedback on the use and acceptability of the SAFS.

Enrolled patients will continue to receive all other services routinely provided by the enrolling Craniofacial Center. Surgeon-raters' use of the SAFS will be assessed, as it would be used in a multi-disciplinary craniofacial team care setting.

**In-Depth Interviews (IDIs).** Each surgeon-rater will have an IDI, one-on-one, by a trained interviewer that will occur within three days from when the surgeon-rater has conducted the SAFS assessment for the patient. These IDIs qualitatively explore the surgeon-rater's initial treatment plan and the decision making process to perform surgery.

The interviewer will be a clinical psychologist with experience and skill in focused discussions in both corporate research and academic settings. The surgeon IDIs will be conducted by telephone.

### 5 STUDY ENROLLMENT AND WITHDRAWAL

A total of 48 patient subjects (32 patients in need of lip revision surgery and 16 patients in need of primary lip repair surgery) will be enrolled. The patients slated for lip revision and lip repair will be enrolled across the six Craniofacial Centers.

#### 5.1 Subject Inclusion Criteria

In order to be eligible to participate in this study, an individual must meet all of the following criteria:

##### 5.1.1 *Lip Revision Subjects*

- Patient has presence of a previously repaired unilateral or bilateral cleft lip and palate with a complete cleft of the primary palate and at least a partial or complete cleft of the secondary palate.
- Patient has a professional clinical recommendation by the craniofacial plastic / oral maxillofacial surgeon for a full or partial thickness lip revision.
- Patient (depending on age) and guardian have ability to comprehend verbal instructions in English, Spanish, or Chinese.
- Patient (depending on age) or guardian able to give consent / assent and has an ability to provide a signed and dated informed consent form in English, Spanish, or Chinese.
- Patient (depending on age) or guardian willing to comply with all study procedures and be available for up to 2 study visits.
- Patient age 4 to 21 years.

##### 5.1.2 *Primary Lip Repair Subjects*

- Patient has presence of an unrepaired unilateral or bilateral cleft lip and palate with a complete cleft of the primary palate and at least a partial or complete cleft of the secondary palate.
- Parent/guardian is willing and able to provide a signed and dated informed consent form in English, Spanish, and Chinese.
- Parent/guardian is willing and able to comprehend verbal instructions in English, Spanish, or Chinese.
- Parent/guardian is willing and able to comply with all study procedures and be available for the duration of the study visits.
- Patient age birth to 8 months.

##### 5.1.3 *Surgeon-Rater Subjects*

The surgeon-raters for this study were previously recruited. Below are the inclusion criteria used for their recruitment.

- A majority of surgeon-raters' cases (>50%) must be patients with the presence of a previously repaired unilateral or bilateral cleft lip and palate with a complete cleft of the primary palate and at least a partial or complete cleft of the secondary palate.
- The surgeon-raters should belong to a Cleft lip and palate team as defined by the ACPA Team standards document (*www.acpa-cpf.org/team_care/*). Each team will have a high volume of patients (> 250 patients per year, primarily prevalent patients and out-patient clinic appointments) that are being followed and cared for to ensure sufficient ability to recruit patients. Each surgeon-rater should have a minimum of 2 lip revision and 2 lip repair surgeries per 12-month period.
- The total number of surgeon-raters represents a broad range of experience specific to the length of time the surgeon-rater has been treating patients with cleft lip and palate and the length of service on a cleft lip and palate team. Ideally, the length of time specific to surgical experience will include the following three categories: Surgeon-raters with < 5 years of experience; 5-10 years of experience, and > 10 years of experience.
- Surgeon-rater willingness to participate and availability for the duration of the study.

#### 5.2 Subject Exclusion Criteria

An individual who meets any of the following criteria will be excluded from participation in this study:

##### 5.2.1 *Lip Revision Subjects*

- Lip revision surgery within the past year.
- A diagnosis of a craniofacial anomaly other than cleft lip (and palate).
- A medical history of collagen vascular disease, or systemic neurologic impairment.
- Mental, visual, or hearing impairment to the extent that comprehension or ability to perform tests associated with the collection of the imaging data is hampered.

##### 5.2.2 *Primary Lip Repair Subjects*

- A diagnosis of a craniofacial anomaly other than cleft lip (and palate).
- A medical diagnosis of collagen vascular disease, and systemic neurologic impairment.

- Mental, visual, or hearing impairment to the extent that the infant's ability to perform tests associated with the collection of the imaging data is hampered.

5.2.3 ***Surgeon-Rater Subjects***

- Surgeons who have previously used the 3D facial images and the dynamic facial movement data of the SAFS protocol prior to their enrollment in this study.

#### 5.3 Strategies for Recruitment and Retention

5.3.1 ***Patients and Guardians***

A limited waiver of HIPAA authorization will be obtained at each center for the identification of potential patient subjects. Patient subjects will be identified by one of the participating plastic surgeons and/or site staff. The surgical schedule from the craniofacial centers and/or plastic surgery departments at MGH, BCH, SHC-BOS, TMC, UNC, and WFB, will be reviewed to identify patients and to determine initial patient eligibility. Potential subjects will be approached by the participating plastic surgeons and/or site staff. During the screening process the site staff will discuss eligibility requirements. No advertising or recruitment material will be used. All potential patient subjects will be contacted directly by the participating plastic surgeons or site staff.

NOTE: Site staff are persons who are employed by the surgery sites and have access to the patient medical record. The participating surgery site will verify that these site staff may access patient medical records for the purpose of this study.

For patient subjects age <18 years all consents will be obtained from the guardians and assent from the patients, as applicable. For patient subjects age ≥18 years all consents will be obtained from the patient.

After screening is completed at the Craniofacial centers, the site staff will contact the Facial Animation Lab Staff (FALS) to review inclusion/exclusion criteria and provide subject contact information.

FALS will contact patients (and their guardian) and schedule an appointment for them to visit one of the Facial Animation Labs. Upon arrival, patients will be given a brief tour of the facility and shown the equipment that will be used for the study. If patients are still willing to participate in the study the FALS will obtain informed consent for the full study.

Data will be collected from the patients on one or two separate pre-surgery visits. The pre-surgery visits will occur at no greater than 3 months before the scheduled surgery, and if needed, a second visit to complete data collection will occur before the surgery for both patient groups.

The guardian or patient, as applicable, will be provided with up to $50.00 to assist with parking at each data-collection visit. In addition, the guardian or patient, as applicable, will be provided with $500.00 at the completion of the total data-collection for the patient even if this requires two visits.

Regular reminders of up-coming appointments for facial animation imaging will be made by the FALS. The surgeon-raters will be reminded of their SAFS assessments on a similar time schedule.

#### 5.4 Treatment Assignment Procedures

##### 5.4.1 *Randomization Procedures*

There will be no randomization procedures. Patients will be approached to participate in the study when they present for treatment. Patients (and their guardians) meeting the selection criteria and willing to consent to participate will be enrolled.

##### 5.4.2 *Masking Procedures*

The researchers, patient subjects, surgeon-raters and site staff at BCH, MGH, SHC-BOS, TMC, WFB, and UNC will not be masked.

#### 5.5 Subject Withdrawal

A guardian may withdraw consent for a minor, and the patient will be withdrawn from the study. A non-minor patient or surgeon-rater may withdraw voluntarily from the study or the investigator may terminate either individual's participation. Should a patient withdraw or be withdrawn by the investigator after informed consent, data collection will be terminated, and demographic information about the patient free from PHI and HIPAA identifiers will be recorded. The reason for withdrawal as well as the time of withdrawal will be collected. The data from these patient subjects may be used for future analyses.

Unless the patient/guardian specifically withdraws consent for the surgeon-rater to perform his/her separate evaluations, those may continue and may be analyzed with the other available study data for the withdrawn patient.

##### 5.5.1 *Reasons for Withdrawal*

Patients and surgeon-raters are free to withdraw from participation in the study at any time.

The study PI may terminate a study participant's participation in the study if:

- Any clinical adverse event (AE) or other medical condition or situation occurs such that continued participation in the study would not be in the best interest of the patient.
- The patient meets an exclusion criterion (either newly developed or not previously recognized) that precludes further study participation.

The study PI may terminate a surgeon-rater's participation in the study if:

- There is a personal emergency on the part of the surgeon-rater and he/she cannot continue participation.
- The surgeon-rater does not comply with the protocol.

##### 5.5.2 *Handling of Subject Withdrawals or Subject Discontinuation of Study Intervention*

###### 5.5.2.1 Patient/Guardian Withdrawal

After the patient (and guardian when applicable) has attended the 1st visit, a 2nd visit will be scheduled, if needed, to complete data collection. The 2nd visit can be scheduled as early as one week after the 1st visit but no later than one day prior to surgery. If a patient/ guardian fails to come to the 2nd visit within the stated timeline, the patient will be withdrawn from the study but his/her data may be used for future analyses. Any patient withdrawn at this stage will be replaced.

###### 5.5.2.2 Surgeon-Rater Withdrawal

A participating surgeon-rater may withdraw from the study at any time. Should a surgeon-rater withdraw from the study, then other surgeon-raters will be screened for their possible participation according to the selection criteria of the surgeon-rater that withdrew (e.g. experience level as detailed in Section 5.1).

#### 5.6 Premature Termination or Suspension of Study

This study may be suspended or prematurely terminated if there is sufficient reasonable cause. Written notification, documenting the reason for study suspension or termination, will be provided by the suspending or terminating party to the PI and the funding agency. If the study is prematurely terminated or suspended, the PI will provide written notification of the reason(s) for termination or suspension to the investigators/centers, oversight committee, IRBs, and Program Official.

Circumstances that may warrant termination include, but are not limited to:

- Determination of unexpected, significant, or unacceptable risk to patient subjects.
- Data that is not sufficiently complete and/or evaluable as determined by the steering committee.

### 6 STUDY INTERVENTION

For each of the surgeon-raters across the six Craniofacial Centers, the SAFS (assessment materials) for four lip revision patients and two primary lip repair patients will be displayed to the surgeon-rater for surgical treatment planning. The surgeon-raters will review the SAFS (assessment materials) for patients who received surgery from another participating surgeon-rater in the study, outside of his/her own clinical center.

The SAFS will be selected from the patients whose data have been collected as part of the study. The patient and surgeon-rater selection process will proceed as follows.

*Patient allocation.* Ideally, recruitment of patients will be approximately balanced across the centers. The two surgeons at MGH also recruit patients from SHC-BOS. Thus, for the purposes of the numbers of patients recruited, MGH and SHC-BOS will be considered one center giving a total of five centers for patient recruitment. Six patients slated for lip revision will be recruited from three of the craniofacial centers and seven patients from the remaining two craniofacial centers. In addition, three patients slated for lip repair will be recruited from four of the craniofacial centers and four patients from the remaining center. A Center, however, may recruit more than 6 to 7 lip revision patients and more than 3 to 4 lip repair patients. Thus, the patient assignment to surgeon-raters will be conducted as follows.

- Surgeons at BCH (Surgeons 1 and 2) will conduct SAFS on patients recruited from Centers MGH/SHC-BOS, TMC, UNC, and WFB only.
- Surgeons at MGH/SHC-BOS (Surgeons 3 and 4) will conduct SAFS on patients recruited from BCH, TMC, UNC, and WFB only.
- Surgeons from UNC (Surgeons 5 and 6) will conduct SAFS on patients from BCH, MGH/SHC-BOS, TMC, and WFB only.
- Surgeons from WFB (Surgeons 7 and 8) will conduct SAFS on patients from BCH, MGH/SHC-BOS, TMC, and UNC only.
- Surgeon from TMC (Surgeon 9) will conduct SAFS on patients from BCH, MGH/SHC-BOS, UNC, and WFB only.

During the study, should any one craniofacial center recruit a maximum of 16 lip revision patients and a maximum of 8 lip repair patients, then the recruitment process will be revised to ensure that enough patients are available for allocation to the surgeon-raters at that Center.

Before the surgeon-rater is notified that the assessment material is available for viewing, the assessment media will be reviewed by the FALS as a quality control measure to ensure standardized formatting. In order to maintain and demonstrate fidelity and competence throughout the study, surgeon-raters and site staff will be trained in the SAFS usage and these procedures will be reviewed annually. In order to ensure compliance with the assessment/SAFS process, surgeon-raters will answer a series of questions during and at the end of each assessment phase described below. Percent and completeness of responses will be recorded for each assessment phase. Dr. Trotman (PI) and Dr. Phillips (co-I) will also review the responses. Trained FALS will ensure that the surgeon-rater views and completes the assessment responses.

#### 6.1 Assessment Phases

For each patient, the surgeon-rater's evaluation will include four phases. At each phase of the evaluation process, the surgeon-rater will be prompted to describe the problem list and surgical goals.

##### 6.1.1 *Facial Form/Disfigurement*

The following phases will assess facial form and disfigurement. Phases 1 and 2 are considered to reflect current practice and are therefore acting as the baseline assessments.

**Phase 1**: This is the surgeon-rater's initial examination of the patient. For this examination, the surgeon-rater is presented with standardized 2D images of the patient.

**Phase 2**: The surgeon-rater will perform a systematic subjective assessment of the patient's static form by viewing and evaluating pre-surgical facial 3D photographs.

Static facial form will be assessed subjectively from a series of standardized facial photographic views and z scores of facial form.

##### 6.1.2 *Facial Impairment*

The following phases will assess facial impairment.

**Phase 3**: The surgeon-rater will perform an assessment of the patient's 'dynamic' facial form by viewing and evaluating pre-surgical videotapes of the patient's animated behaviors.

**Phase 4**: The surgeon-rater will evaluate the dynamic objective measures displayed as visual aids (statistical modeling) and z scores of circumoral movement.[19]

#### 6.2 Surgeon-Rater's Goals/Decision-Making for Surgery

Following each assessment phase (1 through 4), a detailed problem list and a list of desired goals that the surgeon-rater would like to achieve with the surgery will be documented for each patient based on the following:

###### 6.2.1 *Goals based on Static Facial Conditions (Phases 1 & 2)*

For this static evaluation, the surgeon-rater will be given the choices below, but will not be limited to these choices and may add goals as indicated/needed.

1) Improve lip vermillion shape
2) Improve symmetry of lip vermillion
3) Increase or decrease upper lip length
4) Improve shape of nasal alar and/or nasal sill
5) Improve symmetry of nasal alar and/or nasal sill
6) Lengthen columella
7) Improve shape of nasal dorsum.

The surgeon-raters also may comment on goals for movement during these phases based on the static photographs of different animations.

###### 6.2.2 *Goals based on Dynamic Facial Conditions (Phases 3 & 4)*

For this dynamic evaluation, the surgeon-raters will be given the choices below but will not be limited to these choices and may add goals as indicated/needed.

1) Increase or decrease the vertical movement of upper lip
2) Increase or decrease the lateral movement of upper lip
3) Improve the symmetry of upper lip movement
4) No change in upper lip movement indicated, change the lip form only.

The surgeon-raters also may comment on goals from the static phase during these phases based on the dynamic facial conditions.

##### 6.3 Administration of Intervention

For each patient being evaluated, there will be a separate administration of the SAFS to the designated surgeon-rater. After each administration, the surgeon-rater will be permitted additional viewings of the SAFS should he/or she request it. The number of viewings and the type of images viewed will be tracked. For each of the surgeon-raters, there will be four SAFS, one for each of four patients evaluated in the lip revision group to give a total of 32 SAFS administered for the lip revision group; and two SAFS, one for each of the two patients evaluated in the lip repair group to give a total of 16 SAFS administered for the lip repair group. Thus, there will be an overall total of 48 SAFS.

Each SAFS assessment will consist of one patient case selected from a craniofacial center for which the surgeon-rater is unaffiliated. With a study SRA present for assistance, standard 2D facial images will be presented to the surgeon-rater through a computer interface and the surgeon-rater will be asked to describe his/her initial treatment plan. The study SRA then will record these comments. Then the SAFS static facial conditions (3D static facial images and z scores) and dynamic facial conditions (2D facial dynamic videos, 3D dynamic statistical movement comparisons, and z scores) will be presented to the surgeon using the computer interface, and as before, the surgeon will be asked to describe the treatment plan and any additional goals for treatment. The study SRA will record these comments.

Initially, the images will be presented to the surgeon-rater in sequence but the surgeon-rater may return and view any of the images more than once. The number of times each image is viewed will be tracked by the SRA. At the conclusion of viewing the images, an IDI will be conducted with the surgeon-rater either immediately or within 3 days to evaluate how the surgeon-rater's treatment plan and goals for surgery may have changed based on the SAFS images.

##### 6.4 Procedures for Training and Monitoring Intervention Fidelity

Dr. Trotman will train the FALS in the use of the SAFS and in the method of administering the SAFS to the surgeon-raters. Under the supervision of Dr. Trotman, the FALS will review pre-surgery data for 10 lip revision patients that were previously compiled for viewing on a DVD to ensure that the FALS understand the static and dynamic facial features of these patients. Dr. Trotman also will instruct the FALS in the use of the SAFS checklist when the FALS meet with the surgeon-raters to review each patient's treatment plan. Dr. Trotman will conduct refresher training of existing staff as needed, but annually at a minimum.

Prior to their first SAFS review, each surgeon-rater will be trained in the use of the Intervention. Each surgeon-rater will perform 2 mock surgical plans using the pre-surgery data for 2 lip revision patients who were previously tested as part of another study. The patients' SAFS data will be compiled on a password protected flash drive for viewing by the surgeon-raters. The surgeon-raters will view the SAFS for the same three 'training' patients at one sitting and will develop a surgical treatment plan for each patient. The overall approach will include the progressive assessment phases to determine the specific set of problems and subsequent surgical goals that are recorded at each phase in a step-wise manner.

During the course of the study, the surgeon-rater, assisted by the FALS, will view the SAFS assessment and respond to the associated questions. The assessment will not be considered complete until answers are provided for all questions. The surgeon-rater's fidelity will be monitored by a checklist of items that the FALS will confirm that the surgeon-rater has completed. As an additional check, the SAFS assessment sessions will be audio recorded to be reviewed by the FALS for confirmation of completed answers by the surgeon-rater.

##### 6.5 Assessment of Subject Compliance with Study Intervention

###### 6.5.1 *Compliance by Patient Subjects*

Patients must participate in creating a series of facial images for analysis. Before their surgery, they will have 1-2 visits to complete the pre-surgery image generation and data collection. Compliance will be assessed by a checklist for each static or video image that will be completed by the FALS to ensure that the facial images and facial animations as well as the required repetitions of each image and animation are fully completed. As a second level of quality control, after the FALS at one of the two data collection sites (either TUSDM or UNC) completes imaging for a patient, then the FALS from the other data-collection site will review the data collected to ensure accuracy of the original checklist.

##### 6.5.2 *Compliance by Surgeon-Rater Subjects*

Surgeon-raters must complete the SAFS assessment and the In-depth Interviews (IDIs). The SAFS assessment follows a set format with a checklist for each assessment Phase of the SAFS. The PI and the FALS will determine compliance by the surgeon-rater with the completion of the SAFS checklist. The IDIs follow a guide. Compliance with the interviews will be assessed by the interviewer who will determine that all questions on the guide have been answered by the surgeon-rater.

#### 6.6 Study Procedural Intervention(s) Description

The SAFS is comprised of a combination of the following data listed within the sub-sections herein, as presented to and assessed by the surgeon-raters.

##### 6.6.1 *2D Static Photographs*

###### 6.6.1.1 Patients Aged Birth-8 Months

Standardized 2D facial photographic images of the patient's face will be captured using a digital camera. Different animated images will be elicited in response to different taste and smell stimuli, and with the use of toys and rattles. The number of images captured will vary per infant.

###### 6.6.1.2 Patients Aged 4-21 Years

Standardized 2D facial photographic images of the patient's face will be captured during different animation or facial movements that include maximum smile, grimace, mouth open, cheek puff, lip purse, and natural smile using a digital camera.

##### 6.6.2 *3D Static Photographs*

###### 6.6.2.1 Patients Aged Birth-8 Months

Photographic images of the patient's face will be captured using the 3dMD Image Acquisition software. Different animated images will be elicited in response to different taste and smell stimuli, and with the use of toys and rattles. The number of images captured will vary per infant.

###### 6.6.2.2 Patients Aged 4-21 Years

Photographic images are captured of the patient's face during different animation or facial movements that include maximum smile, grimace, mouth open, cheek puff, lip purse, and natural smile using the 3dMD Image Acquisition software.

##### 6.6.3 *Facial Videography*

###### 6.6.3.1 Patients Aged Birth-8 Months

Video of the patient's face will be captured. As with the 3D Static Photographs, different animations will be elicited in response to different taste and smell stimuli, and with the use of toys and rattles.

###### 6.6.3.2 Patients Aged 4-21 Years

As before, video recordings of each patient's face will be captured during different animations or facial movements that include maximum smile, grimace, mouth open, cheek puff, lip purse, and natural smile.

##### 6.6.4 *Facial Movement Data*

###### 6.6.4.1 Patients Aged Birth-8 Months

Facial movement is collected using motion capture systems. The purpose is to measure the movement of the face at specific sites and especially concentrated on the perioral region.

###### 6.6.4.2 Patients Aged 4-21 Years

Facial movement is collected using motion capture systems. The purpose is to measure the movement of the face at specific sites and especially concentrated on the perioral region. The individual is instructed to make different animations or facial movements that include the following six maximum facial animations in the sequence determined by a randomization schedule: grimace, lip purse, cheek puff, big smile, mouth opening, and natural smile.

#### 6.7 Administration of Procedural Intervention

All media on a patient will be presented to the assigned surgeon-rater in the following order, by media category: 1) the 2D static images; 2) the 3D static images + z scores; and 3) the standard 2D videos, and 4) the Cortex Movement Comparison videos + z scores. There will be multiple media files within the 3D static group and the Cortex Movement Comparison group.

Each category of media will be accompanied by a set of associated questions to which the surgeon-rater will respond. The surgeon-rater's assessment of the patient's media need not be completed in a single session; however, the assessment will not be considered complete until the surgeon-rater has answered all questions.

### 7 STUDY SCHEDULE

See Appendix A for Schedule of Events.

#### 7.1 Screening

Patient subjects will be recruited due to planned lip revision/repair surgery by one of the participating plastic or oral maxillofacial surgeon-raters. Once the surgical schedule has been reviewed, the patient/guardian will be approached by site staff to determine interest and eligibility. Then, the following will be conducted:

- Preliminary review of the medical/dental history to determine eligibility based on inclusion/exclusion criteria.
- Review medical records in order to confirm eligibility.

Craniofacial Center site staff will provide contact information for the patient to the FALS. The FALS then will contact the patient to schedule the study visit, and provide potential patients with instructions needed to prepare for the study visit.

#### 7.2 Patient Enrollment

##### 7.2.1 *Visit 1*

- Obtain and document assent/consent from patient/guardian.
- Verify inclusion/exclusion criteria.
- Obtain demographic information, and review and confirm medical/dental history.
- Obtain the 2D and 3D static images and video and collect data for the SAFS.
    - Collect static 2D facial data of the patient.
    - Collect static 3D facial data of the patient.
    - Collect 2D video data of the patient's face during different facial animations.
    - Collect 3D dynamic movement data of the patient's face during different animations.
- Record patient's compliance with obtaining the different images that comprise the media portion of the SAFS.
- Record any UPs as reported by patient or observed by investigator.

##### 7.2.2 *Visit 2*

- Collect data not completed during Visit 1.
    - Collect static 2D facial data of the patient.
    - Collect static 3D facial data of the patient.
    - Collect 2D video data of the patient's face during different facial animations.
    - Collect 3D dynamic movement data of the patient's face during different animations.
- Record patient's compliance with the obtaining the different tests that comprise the media portion of the SAFS.
- Record any UPs as reported by patient or observed by investigator.

##### 7.2.3 *Withdrawal Visit*

There will be no patient withdrawal visit. If a patient/parent withdraws, the decision will be documented along with a reason for the withdrawal.

##### 7.2.4 *Unscheduled Visit*

NA

#### 7.3 Surgeon-Rater Study Schedule

##### 7.3.1 *Intervention and Decision Visit*

###### 7.3.1.1 SAFS Intervention

A member of the study team will administer a patient's SAFS to the surgeon-rater in person.

- Administer the patient's SAFS to the surgeon-rater.
    - Surgeon-rater performs the static 2D facial photographic evaluation of the patient.
    - Surgeon-rater performs the static 3D facial photographic evaluation of the patient.
    - Surgeon-rater performs the 2D dynamic videography facial evaluation of the patient.

- Surgeon-rater performs the 3D dynamic movement, statistical-comparison evaluation of the patient.

##### 7.3.2 *Surgeon-Rater's Compliance*

Record surgeon-rater's compliance with the different evaluation phases that comprise the Intervention using the Intervention checklist.

##### 7.3.3 *In-Depth Interview (IDI)*

An interview will be conducted via telephone either immediately or within 3 days of a surgeon-rater's participation in a patient's SAFS. If there is a delay prior to the IDI, the surgeon-raters will be presented with the output of their SAFS in order to prepare for the IDI

- IDI conducted with the surgeon-rater to discuss his/her decision making process to perform surgery and the initial treatment plan.
- Record surgeon-rater's compliance with the IDI as assessed by the interviewer using the interview guide.

### 8 STUDY PROCEDURES / EVALUATIONS

#### 8.1 Testing for Allergies in All Patients

One of the study procedures involves the placement of small markers on the patient's face. For the child patient, these markers are secured to the face with a non-latex based eyelash adhesive. For the infants, either non-latex adhesive or double-sided surgical adhesive tape will be used. All patients will be tested prior to placement of the markers by placing a small amount of eyelash adhesive on the patient's finger, back of the hand and/or the upper arm to ensure that there is no allergy. Should a patient display any signs of possible allergy, the area will be treated with 1% hydrocortisone cream and double-sided surgical adhesive tape will be used to secure the markers.

#### 8.2 Image Data Collection for Subjects Aged 4-21 Years

This section outlines the specific data collection imaging methods for subjects aged 4 to 21 years only.

##### 8.2.1 *2D Static Photographs*

The patient will be seated comfortably at a set distance in front of a digital camera and instructed to relax and focus on the camera. Then several different animations will be photographed that include rest, big smile, natural smile, lip purse, cheek puff, grimace, and mouth opening. The order of capturing the photographs will be randomized at the time that the patient ID is assigned. Once all facial photographs have been captured, they will be uploaded for review by the surgeon-raters.

##### 8.2.2 *3D Static Photographs*

The patient will be seated comfortably in front of the camera and instructed to relax and focus on an object at a set distance. Then several different animations will be photographed that include rest, big smile, natural smile, lip purse, cheek puff, grimace, and mouth opening. The order of capturing the photographs will be randomized at the time that the patient ID is assigned. Once all photographs have been captured, they will be uploaded for review by the surgeon-raters.

##### 8.2.3 *2D Facial Videography*

The participant is seated comfortably, and the cameras are positioned to record the frontal with both profile views of the patient's face. The participant's face fills each camera view within the video monitor. Each animation is demonstrated to the participant and the participant is asked to mimic the movement in order to ensure that s(he) is familiar with this movement. The participant is instructed to make different animations using a randomized sequence that include: maximum smile, natural smile, mouth open, cheek puff, lip purse, and grimace. The animations will be videotaped, and processed for viewing by surgeon-raters.

##### 8.2.4 *Movement Analysis Systems (Motion Analysis™ & 3dMD Dynamic System™)*

Facial movement is collected at both TUSDM and UNC using a marker-based Motion Analysis camera system. The data from the Motion Analysis Camera will be used for the SAFS. For exploratory analyses only, facial movement also will be collected using a second markerless 3dMD dynamic system camera at the TUSDM site only. The data from the 3dMD dynamic system camera will not be part of the SAFS.

(a) Motion Analysis™ System

The purpose is to measure the movement of the face at specific sites especially concentrated on the perioral region. At each image data-collection visit, the patient will perform a series of repeated facial animations that will be recorded using a motion analysis system.

The FALS will secure markers to specified sites on the patient's face with either eyelash adhesive or double adhesive tape. The patient will be seated comfortably in front of the cameras. The animations will be demonstrated to the patient, and the patient will mimic the movement in order to ensure that the markers are firmly attached to the patient's skin (i.e., no slippage) and that s(he) is familiar with the different movements. After several practice trials, the patient will be instructed to make different facial animations or movements that include the following five maximum facial animations: lip purse, cheek puff, big smile, mouth opening, and grimace. Each of the animations will be repeated up to 10 times. All animations are randomized except the grimace, which is always recorded last. Prior to recording the grimace, two markers are removed at the base of the nose to obtain good data. Once the recording session is completed, any residual adhesive will be gently removed from the participant's face using a gauze pad with distilled water. The data then will be processed for viewing by surgeon-raters.

(b) 3dMD™ Dynamic System

The patient will be seated comfortably in front of the cameras. The patient will be instructed to make different facial animations or movements that include up to five maximum facial animations: lip purse, cheek puff, big smile, mouth opening, and grimace. Each of the animations will be repeated up to 3 times. All animations are randomized. The data then will be processed for viewing by surgeons.

#### 8.3 Image Data Collection for Subjects Aged Birth-8 months

This section outlines the specific data collection methods for infants (birth-8 months of age) only.

##### 8.3.1 *2D Static Photographs*

The infant is seated comfortably on the caregiver's lap at a set distance in front of a digital camera and the parent holds the infant firmly on his/her lap. The parent and infant are properly positioned so that the infant is centered within the frame of view of the camera. By means of toys, rattles, and other aids, photographic images of the infant's face are captured at rest, smiling, and crying. The number of images captured will vary per infant. Some infants will not cry or smile, but substantial efforts should be made to elicit these movements.

###### 8.3.2 *3D Static Photographs*

The infant is seated comfortably on the parent's lap in front of the camera and the parent holds the infant firmly on his/her lap. The parent and infant are properly positioned so that the infant is centered within the frame of view of the camera. By means of toys, rattles, and other aids, photographic images of the infant's face are captured at rest, smiling, and crying using the 3dMD Image Acquisition software. The number of images captured will vary per infant. Some infants will not cry or smile, but substantial efforts should be made to elicit these movements.

###### 8.3.3 *2D Facial Videography*

The video cameras are positioned to record the frontal and lateral views of the infant's face and the infant's face is positioned to fill the view for each camera on the video monitor with enough room for sudden movements. Then, the infant's movements are recorded for approximately 15 minutes, capturing all 3 views simultaneously. During the recording, smiles, cries, and other facial animations will be elicited. Also, the caregiver will be asked to administer a small amount of taste stimuli solutions (described in Section 8.3.4) to elicit movements.

###### 8.3.4 *Movement Analysis System (Motion Analysis™ & 3dMD Dynamic System™)*

While the parent holds the infant, 14 markers will be placed on the infant's face as specified by the templates. The infant is secured in a modified car seat and positioned with the face towards the cameras. During the recording session, different animations will be elicited using toys and rattles. The desired spontaneous movements include: smiles, crying, suckling, and taste responses. In addition, movements will be elicited in response to various taste stimuli. Infants will be given a small amount of sucrose, sodium chloride, and citric acid solutions by mouth using a disposable plastic syringe, and the facial movements will be recorded. Between stimuli solutions, the infant's mouth will be rinsed with filtered water.

##### 8.4 Surgeon-Rater Assessment Method/Standardized Protocol

Once the performing/operating surgeon and patient/guardian have discussed the possibility of surgery, those patients/guardians who consent to participate in the study will be scheduled to travel to the closest study site for data collection. The data will be used to compile the SAFS and the assigned surgeon-rater will use the SAFS to guide decisions and develop a hypothetical surgical treatment plan. The overall approach will include progressive assessment phases to determine the specific set of problems and subsequent surgical goals that will be recorded at each phase in a step-wise manner (see the Manual of Procedures (MOP) for details).

#### 8.5 Surgeon-Rater In-Depth Interviews (IDIs)

Each surgeon-rater will participate in structured in-depth telephone interviews (IDIs) designed to elicit information on treatment planning decisions as follows:

(a) For each of the surgeon-rater, the interviewer will conduct an IDI after the SAFS for four lip revision and two lip repair patients to give a total of 48 IDIs.

Unless saturation is reached, as determined by Dr. Trotman (PI), and depending on surgeon-rater availability, a target number of up to 10 IDIs will be conducted per quarter (every 3 months).

##### 8.5.1 *Surgeon-Rater In-Depth Interview*

Prior to the SAFS being administered to the surgeon-rater, the FALS at each testing site will schedule the IDI to be conducted within **3 days of completion** of the SAFS.

- The images that comprise the SAFS will be made available to the surgeon-rater to review as needed.
- The IDI will take approximately one hour and will be conducted by the interviewer via telephone.
- The interviewer will follow a pre-surgery IDI guide that focuses on the surgeon-rater's thoughts and plans concerning surgery; how the SAFS affected the surgeon-rater's thoughts and plans; the extent of information about the surgery that they would provide to the patient/guardian and why; extent of information about the treatment, including expectations and goals for surgery; and a discussion on the drivers for the final decision to perform surgery.
- The surgeon-rater will be asked to answer all questions posed by the interviewer who will follow an interview guide.
- The interviewer will record details of the entire interview.
- The interviewer will conclude the interview and answer any questions from the surgeon-rater.
- All interviews will be recorded for subsequent transcription and coding.

The interview guide will be modified during the project period based on surgeon-raters' responses. The general format for the interview guides will be inclusive but designed to elicit open-ended discussion, and there will be separate guides for the infant and non-infant patients.

### 9 ASSESSMENT OF SAFETY

#### 9.1 Specification of Safety Parameters

Lip revision and primary lip repair surgeries for patients with CL/P are standard of care. Adverse events may occur as a result of the study procedures or as a result of the surgery.

Because patient participation in this study ends prior to surgery, and because the patient-specific SAFS occurs on a surgeon-rater who is not the patient's surgeon, only information related to unanticipated problems will be collected.

##### 9.1.1 *Unanticipated Problems*

The Office for Human Research Protections (OHRP) considers unanticipated problems involving risks to subjects or others to include, in general, any incident, experience, or outcome that meets **all** of the following criteria:

- unexpected in terms of nature, severity, or frequency given (a) the research procedures that are described in the protocol-related documents, such as the IRB-approved research protocol and informed consent document; and (b) the characteristics of the subject population being studied;
- related or possibly related to participation in the research ("possibly related" means there is a reasonable possibility that the incident, experience, or outcome may have been caused by the procedures involved in the research); and
- suggests that the research places subjects or others at a greater risk of harm (including physical, psychological, economic, or social harm) than was previously known or recognized.

##### 9.1.2 *Adverse Events and Serious Adverse Events*

Data on adverse events and serious adverse events will not be recorded or reported except as they qualify as an unanticipated problem (see Section 9.1.1),

#### 9.2 Halting Rules

There are no formal halting rules in this study.

#### 10 STUDY OVERSIGHT

In addition to the PI's responsibility for oversight, study oversight will be under the direction of a Medical Monitor. The PI will prepare an annual report summarizing unanticipated problems (UPs), study conduct, and progress. This report will be reviewed by the Medical Monitor, who will provide recommendations to the NIDCR. If major concerns arise, more frequent reports may be requested.

The PI and PO will also communicate approximately quarterly to assess safety and efficacy data (if applicable), study progress, and data integrity for the study. In the unlikely event that safety concerns arise, more frequent meetings may be held.

#### 11 CLINICAL SITE MONITORING

Clinical site monitoring will not be done for this study; however, the NIDCR reserves the right to conduct independent audits or clinical monitoring as necessary.

### 12 STATISTICAL CONSIDERATIONS

This research is designed to investigate the following areas: Understanding how surgeon-raters incorporate the SAFS into their practice and decision-making processes (study objectives 1 and 2); quantitatively assessing the extent to which the SAFS changes surgeon-raters' problem list and treatment planning goals for lip revision (study objective 3); quantitatively assessing the extent to which the SAFS changes surgeon-raters' problem list and treatment planning goals for primary lip repair (study objective 4); and quantitatively assessing the extent to which the SAFS changes surgeon-raters' problem list and treatment planning goals for lip revision as a function of surgical expertise (study objective 5);

#### 12.1 Study Hypotheses

We hypothesize the following.

For study objectives 1 and 2:

$H_0$: With information collected during the IDIs, no common themes will be identified for surgical care of patients for either lip revision or for primary lip repair.

For study objectives 3 and 4:

$H_0$: The additional information from the animated videos and objective measures/visual aids of the SAFS will not produce a change in the surgeons' problem list or treatment goal for a majority of lip revision and primary lip repair patients (>70%).

For study objective 5:

$H_0$: Surgeon-raters with less clinical experience/time in practice versus those with greater clinical experience find that the objective measures of the SAFS are not helpful for treatment planning for lip revision surgery versus the subjective assessment of the SAFS.

#### 12.2 Sample Size Considerations

##### 12.2.1 *Precision Estimates / Sample Size for Objectives 1 and 2 (Qualitative Analysis).*

The study was powered on the primary outcome measures related to the qualitative data analysis. Specifically, qualitative data analysis involves the systematic interpretation and analysis of patterns and themes in verbal (textual) data from the surgeon-raters' IDIs. Ultimately, the goal of qualitative analysis is to make sense of complex narrative or interview data so that the relevant research questions can be answered. Sample size in qualitative research is set at the point when the data collected reveals no new concepts or patterns (termed-theoretical saturation).[26-27] There is no clear consensus on appropriate sample sizes in qualitative research, however, evidence suggests that saturation generally occurs between 10 and 30 interviews.[26,28] Given the different levels of clinical experience of the surgeon-raters, the timing of the IDIs to occur after the SAFS and the planned evaluation of the SAFS for both the lip revision and lip repair groups, we are confident that the total of 48 IDIs to be conducted over a 23 month period will be sufficient to gain insight into surgeon-raters' assimilation and use of the SAFS, and 'learning' effects.

#### 12.3 Final Analysis Plan

##### 12.3.1 *Analysis Plan for Objectives 1 and 2*

Objectives: To qualitatively assess how surgeon-raters integrate the Intervention's objective measures and visual aids with the Intervention's systematic subjective assessment in the decision-making process for the clinical surgical procedures of lip revision or lip repair.

###### 12.3.1.1 Analysis of IDIs and Structured Interviews

IDIs will be conducted with the surgeon-raters after the administration of the SAFS.

All interviews will be recorded and transcribed. The interviewer and Dr. Trotman will review these transcriptions. The general format for the interview guides will be inclusive but designed to elicit open-ended discussion, and there will be separate guides for infants and non-infants. The areas to be explored are modeled on those identified by surgeons involved in the development of the SAFS. The interview guides will focus on how the SAFS affected the surgeon-rater's thoughts and plans concerning surgery. These guides will be modified during the project period based on surgeon-raters' responses.

The analysis will be informed by grounded theory [27, 29-30] which allows for the induction of concepts and understanding from the data as well as for both intended and unintended results. The approach uses existing theory to guide the research with an aim of extending and improving it (see Burawoy's "extended case method").[31-32] Consonant with the principles of grounded theory, we will first code interviews. An iterative process of analysis will be followed in which two members of the research team, assisted by the interviewer and Dr. Trotman, will independently read and discuss a randomly selected subset of IDIs collected at each visit to develop an initial set of codes that reflect a provisional conception of factors likely to be of theoretical importance in the analysis. These codes then will be refined and secondary codes established to clarify and contextualize important themes. Together, these codes will constitute the coding manual. As coding progresses, theoretical memos will be written to reflect on and make sense of emerging concepts and themes in order to move the analysis from surgeon-raters' individual experiences with the SAFS to a higher analytical understanding of the SAFS as a process that integrates, develops, and focuses decisions on treatment need. Early theoretical memos function to organize data and uncover emerging themes. In order to capture surgeon-rater differences, early theoretical memos will be coded by Dr. Trotman to ensure that differences are determined during data reduction. As data analysis progresses, memos become more specific, and narrow the focus of the analysis toward important themes. Once identified, the "constant comparison" method[29] then will be used to define these themes by systematically comparing chunks of data. These comparisons will be analyzed in theoretical memos which will become the basis for the study's findings.[30]

The focus of the analysis is on the integration of the SAFS by surgeon-raters as well as on decision-making. We also will use the data to examine surgeon-rater characteristics and trends over time within and across surgeon-raters, e.g., evaluate how time in practice and training affects the use and integration of the SAFS by surgeon-raters. Since the late 1980s, software programs have evolved to specifically meet the needs of qualitative researchers.[33-34] Programs such as MAXQDA or ATLAS.ti are built on classic qualitative data analytic methods such as grounded theory, discourse analysis, and qualitative content analysis. Such software does not replace the role of the researcher in the interpretation and synthesis of data, but assists in labeling, categorizing, and sorting large amounts of interview data. For the present study, we will use these software packages to analyze qualitative data from surgeon-rater IDIs. In order to establish inter-rater reliability, the two team members will independently review and code randomly-selected interviews, and then come together at regular intervals to discuss codes, achieve consensus, and resolve differences under the supervision of the interviewer and Dr. Trotman. This analytical triangulation will "provide an important check on selective perception and interpretive bias" thereby improving the reliability and validity of our findings.[35-37]

The above will be conducted on the data from patients requiring lip revision to address objective 1 and on the data about patients requiring primary lip repair to address objective 2.

###### 12.3.1.2 Measures/Analyses for the SAFS

The SAFS is made up of objective measures of patient's facial movement (with visual aids) and static form along a systematic subjective assessment of the patient's facial movement and form. The information provided below describes how the objective measures of movement and form are generated for the SAFS.

###### *12.3.1.2.1 3D Objective Facial Movement*

Information presented to surgeon-raters will be dynamic graphics (movies) and numerical measures that represent the difference in the facial movement of the subjects from the non-cleft comparators (data obtained during R01 DE 013814). A motion analysis system will be used to measure the circumoral movements of each subject according to the previous methods and analyses of Trotman and co-workers[1,3]. The system tracks retro-reflective markers secured to specific facial (nasolabial) landmarks, and will be used to capture the kinematic data from both the infant and children patients during a series of set facial animations and behaviors. For each patient, up to 10 replications of each of 6 animations will be obtained. For each facial landmark (approx. 38), a time series of 3-D vectors defined by (x, y, z) are recorded where x, y and z represent position in space at 1/60 second intervals for 4 seconds. Measures of lip function/movement are generated based on the change in distance between landmarks during facial emotive movements, and for each maximum animation (smile, lip purse, cheek puff, grimace, mouth opening) and natural smile. Subsequently, a *statistical* visual modeling comparison is generated of the patients mean movements compared with the mean 'non-cleft' comparator movement for surgeon-raters to view. More specifically, the steps in the data analysis are:

1. For each recorded motion, we check the data for missing values and motion capture errors. Short sequences of missing values are imputed and robust smoothing methods are used to remove rare physical impossible 'glitches' in the recorded motion. In a few cases, the motion will be unusable and will be discarded. Concrete procedures for this stage of the analysis have been established from the previous study and will be used uniformly on the new data.
2. We use the first ten frames of the data to establish the "at rest" position of the face. We consider the collection of 38x3 matrices of 240 shapes (one for each frame) and apply generalized Procrustes analysis to find the first principal component (PC). We locate the frame corresponding to the time at which the first PC is most different from the established rest position. We use the shape formed by the mean plus first PC at this maximal frame as the reported maximum position of the face for this motion.
3. We compute the vector in shape space from the initial position to the maximal position as a representation of the motion.
4. For the ten replications of each motion, we average the motion vectors to form an average motion for the subject/animation combination.

We are now able to construct movies that represent the motion of the subject relative to the control population. From the previous study, we have established the mean at rest shape together with a vector expressing the motion to the maximal pose. We add the average motion vector for the subject to the mean at rest position for the controls. We use this to construct a movie which shows the difference in the motion of the subject relative to the control mean motion.

In addition, we will present a numerical measure representing how different the motion of the subject is relative to the control population. We will compute the Mahalanobis distance of the subject from the control population mean. We will present this score in percentile form for easier interpretation by the surgeon-rater.

The statistical procedures to compute these measures have been developed from previous studies and have been programmed into concrete procedures that will be applied uniformly throughout the study.

*12.3.1.2.2 3D Measures of Static Facial Form*

The 3dMD system will be used to capture pre-specified landmarks from the face. We established a database of control subjects which will be used to generate the baseline distributions to which the subjects will be compared. The subjects will be measured with same system and a Mahalanobis distance of a similar type to the dynamic case will be computed. The score will be presented on a percentile scale for ease of interpretation.

##### 12.3.2 ***Analysis Plan for Objectives 3 and 4***

Objectives: Determine the extent to which the SAFS changes the surgeon-rater's problem list and goals for lip revision and lip repair.

On a general level, the desired goals may change in the following 3 ways:

1) The surgeon-rater may decide not to recommend surgery, so the original desired goals are never achieved;

2) The surgeon-rater may decide to modify the surgery to achieve certain specific goals as detailed in 1 through 7 for the Static Facial Conditions and 1 through 4 for the Dynamic Facial Conditions, so the original desired goals are changed;

   and

3) The surgeon-rater recommends proceeding with the surgery as planned, so the original desired goals are maintained.

The surgeon-rater will be asked for his/her goal(s) for surgery after each phase of the SAFS has been presented.

Any decision in which the surgeon-rater does not recommend the surgery or changes the original desired goal as detailed in a) through d) is counted as 'a change' by the surgeon-rater and coded as '1' for that patient. If the surgeon-rater recommends proceeding with the original plan, this decision will be counted as 'no change' and coded as '0'. Thus, the outcome for this phase will be '0' (no change) and '1' (a change).

###### 12.3.2.1 <u>Sample size estimates</u>

The primary outcome measure that will be assessed is the extent to which the 3D images and objective measures of distortion changes the original targeted goals of the surgeon-rater. The sample size calculation is based on an exact binomial test with a 0.05 one-sided significance level. This test will have 92% power to detect the difference between $H_0$: $p = 0$ and $H_1$ $p = 0.10$ when the sample size is 25.

We will estimate whether the proportions of change across phases 1 to 4 of the SAFS differ from one another as a first step in assessing the relative impact of the Phase-specific components of the SAFS. Specifically, a Mantel-Haensel test for repeated measures[38] will be used with each patient serving as his/her own stratum to assess the association of Phase and change ("yes" or "no"). Separate tests will be conducted for the primary lip repair and lip revision patients.

In addition, to determine in more detail the relative impact of the components of the SAFS, that is the systematic qualitative assessment made by the surgeon-rater versus objective measures, we will determine whether the facial attributes that the surgeon-rater identifies as a problem for a patient is in agreement with objective measures of the area. Separate comparisons of the problem list will be made for the facial form and the facial impairment evaluation. For example, after the surgeon-rater has completed the evaluation of the patient using the SAFS, the results of the qualitative assessment in phases 1 and 2 of the SAFS for a patient's facial form/disfigurement evaluation will be compared with objective measurements for the area. Therefore, should the surgeon-rater determine lip vermillion symmetry and nasal alar shape must be improved, we will assess the agreement between this qualitative determination by the surgeon and objective measurements of these same areas. Likewise, the qualitative results made by the surgeon after viewing the facial video images for a patient's facial impairment in phase 3 of the SAFS will be compared with the objective dynamic measurements of areas (phase 4). Therefore, should the surgeon-rater determine upper lip movement needs to decrease and movement symmetry improved, we will assess the agreement between this qualitative determination by the surgeon and the objective dynamic measurements of these areas. Temporal changes for this agreement will be measured to understand the effects of learning on surgeon-rater integration of the SAFS. For example, over time and with continued use of the SAFS, surgeon-raters may find they tend to diagnose certain problems from the subjective assessment only, as the objective evaluation may serve only to confirm their findings.

###### 12.3.2.2 Statistics for agreement between surgeon-raters' subjective evaluation and the objective measures

For individual facial attributes, bivariate logistic regression[39] will be used to simultaneously model two correlated binary outcomes—the systematic qualitative assessment made by the surgeon-rater and the corresponding objective measure. In addition to specifying a logistic model for each outcome, a third equation will be included in the joint model to estimate agreement between the two binary outcomes, which will be summarized with kappa statistics.[40] The model for kappa will include as a predictor the number of prior SAFSs for the surgeon-rater (equivalently, the sequence number of the surgeon's patient). Surgeon-raters will be included in the model as a fixed effect since their number is small. These surgeon-rater effects will be examined for dependency of agreement on surgeon-rater experience.

##### 12.3.3 ***Analysis Plan for Objective 5***

Objective: Assessment of the extent to which the SAFS changes surgeon-raters' problem list and treatment planning goals for lip revision as a function of surgical expertise.

As stated above, we will determine whether the facial attributes that the surgeon-rater identifies as a problem for a patient is in agreement with the objective measures. Separate comparisons of the problem list will be made for the facial form and the facial impairment evaluation. For example, after the surgeon has completed the evaluation of the patient using the SAFS, the results of the qualitative assessment in phases 1 and 2 of the SAFS for a patient's facial form/disfigurement evaluation will be compared with objective measurements for the area. Therefore, should the surgeon-rater determine lip vermillion symmetry and nasal alar shape must be improved, we will assess the agreement between this qualitative determination by the surgeon-rater and objective measurements of these same areas. Likewise, the qualitative results made by the surgeon-rater after viewing the facial video images for a patient's facial impairment in phase 3 of the SAFS will be compared with the objective dynamic measurements of areas (phase 4). Therefore, should the surgeon-rater determine upper lip movement needs to decrease and movement symmetry improved, we will assess the agreement between this qualitative determination by the surgeon-rater and the objective dynamic measurements of these areas. Comparisons of this agreement will be made among surgeon-raters based on surgeon-rater level of experience (years in clinical practice). For example, surgeon-raters with more years in clinical practice versus those with less years will demonstrate greater agreement between the qualitative and objective measures.

###### 12.3.3.1 <u>3D Objective Facial Movement</u>

Specifically, the motion data will be pre-processed as described previously. We will have measures of the magnitude and symmetry of the motion for specified marker subsets corresponding to the areas of interest in addition to measures of the entire motion of the subject. Also, side-to-side reflection of the face will be used to generate measures of symmetry in the motion for specified subsets of markers of the face. Thus, the surgeon-raters will identify the presence or absence of a problem (and magnitude where appropriate) for these combinations. A set of paired scores will be generated where each pair consists of a surgeon-rater score and an objective score. Standard statistical methods then will be used to rank the degree of agreement and assess how this varies with respect to surgeon-rater level of experience.

###### 12.3.3.2 <u>3D Static Facial Form</u>

Specifically, the statistical procedures used here will be similar to those used for the dynamic comparison. A different set of markers is used so the subsets will be somewhat different and the comparisons will be based on the 'at rest' position rather than the maximal motion.

##### 12.3.4 *Checking Assumptions for Dynamic Measures*

For shape statistics, there is a limited selection of diagnostics available to check assumptions. Even so, we have the advantage of the visualizations of movement to back up the purely numerical analysis. This gives us a reliable way of detecting outliers or other errors due the measurement process.

### 13 SOURCE DOCUMENTS AND ACCESS TO SOURCE DATA/DOCUMENTS

Each participating site will maintain appropriate medical and research records for this study, in compliance with ICH E6, Section 4.9 and regulatory and institutional requirements for the protection of confidentiality of subjects. Each site will permit authorized representatives of NIDCR and regulatory agencies to examine (and when required by applicable law, to copy) research records for the purposes of quality assurance reviews, audits, and evaluation of the study safety, progress and data validity. Source data are all information, original records, observations, or other activities in a clinical study necessary for the reconstruction and evaluation of the study.

### 14 QUALITY CONTROL AND QUALITY ASSURANCE

All site level quality management efforts will be supervised by the site Principal Investigators, aided by the site Study Coordinators and with input from the study PI. Study-level quality oversight will be provided by the study PI. This includes training and oversight of the clinical sites and oversight of the data coordinating center and facial animation lab. Details of the quality processes will be included in the quality management plan.

The Medical Monitor provides quality assessment by reviewing the scheduled reports of aggregated data provided by the study team.

### 15 ETHICS/PROTECTION OF HUMAN SUBJECTS

#### 15.1 Ethical Standard

The investigator will ensure that this study is conducted in full conformity with the principles set forth in The Belmont Report: Ethical Principles and Guidelines for the Protection of Human Subjects of Research, as drafted by the US National Commission for the Protection of Human Subjects of Biomedical and Behavioral Research (April 18, 1979) and codified in 45 CFR Part 46 and/or the ICH E6.

#### 15.2 Institutional Review Board

The protocol, informed consent form(s), recruitment materials, and all patient participant materials will be submitted to the IRB at each site for review and approval. Approval of both the protocol and the consent forms must be obtained before any patient participant is enrolled. Any amendment to the protocol will require review and approval by the IRB at each site before the changes are implemented in the study.

#### 15.3 Informed Consent Process

Each participating institution will be provided with model informed consent and assent forms. Each institution may revise or add information to comply with institution consent templates, but may not remove procedural or risk content from the model consent form. Each institution may also have their consent/assent translated into additional languages depending on their population needs. The consent process will be conducted in the language of the consent form by an appropriate staff member.

Informed consent is a process that is initiated prior to the individual agreeing to participate in the study and continues throughout study participation. Extensive discussion of risks and possible benefits of study participation will be provided to patients and their guardians[1], if applicable. A consent form and assent form, as applicable, describing in detail the study procedures and risks will be given to the patients/guardians. Consent/assent forms will be IRB-approved, and the patients/guardians are required to read and review the documents or have the documents read to him or her. The investigator or designee will explain the research study to the patient/guardian and answer any questions that may arise. The patient/guardian will sign the informed consent/assent document(s) prior to any study-related assessments or procedures. Patients will be given the opportunity to discuss the study with their surrogates or think about it prior to agreeing to participate. They may withdraw consent at any time throughout the course of the study. A copy of the signed informed consent/assent documents will be given to patients/guardians for their records. The rights and welfare of

[1] The term guardian is being used as the generalized term for responsible adult with signatory power for a minor patient. This could be a parent or other legally responsible person.

the patient subjects will be protected by emphasizing to them that the quality of their clinical care will not be adversely affected if they decline to participate in this study.

The consent process will be documented in the clinical or research record.

Surgeon-raters will be consented, unless otherwise decided by their local IRB.

##### 15.4 Exclusion of Women, Minorities, and Children (Special Populations)

Individuals of any gender or racial/ethnic group may participate in this study; however, because of the need for primary lip repair and lip revision surgery within certain age ranges, the study is limited to patients from birth to 21 years.

##### 15.5 Subject Confidentiality

Participant confidentiality is strictly held in trust by the investigators, study staff, and the sponsor(s) and their agents.

All records will be kept confidential. Each patient and surgeon-rater will be assigned a unique ID, which will be used identify that individuals data record and which will ensure confidentiality. The code linking the patient's and surgeon-rater's name and ID number will be securely maintained and only the study staff and investigators will be allowed to access this code. No patient or surgeon-rater will be identified in any reports and publications. In the event that the photographs or video images of a patient are used for educational or presentation purposes, additional consent will be first obtained. All videotapes will be stored in a locked cabinet in a secure room. Only study staff and investigators will have access to the tapes.

The usage of private health information (PHI) in the form of full-face images is required based on the objectives of this study. The study also requires the usage of medical history to verify patient participant eligibility. No PHI will be used without proper consent, and every effort will be taken to ensure that every patient's confidentiality is maintained. No Certificate of Confidentiality will be required for this study.

The study protocol, documentation, data, and all other information generated will be held in strict confidence. No information concerning the study or the data will be released to any unauthorized third party without prior written approval of the sponsor.

The study monitor or other authorized representatives of the sponsor may inspect all study documents and records required to be maintained by the investigator, including but not limited to, medical records (office, clinic, or hospital) for the study subjects. The clinical study site will permit access to such records.

##### 15.6 Future Use of Identifiable Data

It is the intent of the investigators to maintain the patient images after the study is complete. Thus, when a patient and/or parent consents to participate in the study, they will be asked to sign a separate consent form to allow their data, including the facial images and video-recordings, to be maintained after the study has been completed and to be used for future studies. The names and other identifying data will be removed from the image files and coded. Also, identifying information will be redacted from audio recording transcripts. All data will be maintained at TUSDM on secured servers that are password protected. These data will be maintained for seven years after the completion of the study. TUSDM IRB will review future studies, and protections of confidentiality for any future studies with the stored data. Should the patient/parent choose not to have their data maintained, the patient's data files will be purged from the database no later than 2 years after the completion of the study. This length of time will ensure the full completion of all studies planned in the original grant submission.

### 16 DATA HANDLING AND RECORD KEEPING

The investigators are responsible for ensuring the accuracy, completeness, legibility, and timeliness of the data reported. All source documents should be completed in a neat, legible manner to ensure accurate interpretation of data. The investigators will maintain adequate case histories of study subjects, including accurate case report forms (CRFs), and source documentation.

#### 16.1 Data Management Responsibilities

Data collection and accurate documentation are the responsibility of the site staff under the supervision of the investigator. All source documents must be reviewed by the site staff and data entry staff, who will ensure that they are accurate and complete. Unanticipated problems must be reviewed by the investigator or designee.

TUSDM will serve as the DCC for this study and will be responsible for data management, as well as for quality review, analysis, and reporting of the study data.

#### 16.2 Data Capture Methods

Study staff will complete eCRFs via a web-based EDC system hosted by the DCC. Access to the EDC system is password-protected and compliant with 21 CFR Part 11. A unique subject identification number will be used on the eCRFs and in the study database. A subject code log that links patient names to identification numbers will be maintained by the investigators in a secure location. Study data will be transmitted to the DCC via a secure, encrypted Internet connection and stored on a secure server. The EDC system includes internal quality checks, such as automatic range checks, to identify data that appear inconsistent, incomplete, or inaccurate.

Staff at the Facial Animation Labs (UNC and TUSDM) will collect facial imaging data using 3D static photographs and videography. Facial movement data will be captured using a Motion Analysis System. All surgeon interviews conducted by the study psychologist will be recorded. These media files will be stored in password-protected and HIPAA compliant cloud storage.

#### 16.3 Types of Data

Data for this study will include safety information (e.g., adverse events, unanticipated problems), imaging data, and outcome measures (e.g., surgeon interviews).

#### 16.4 Schedule and Content of Reports

Study progress reports will be generated periodically to include information such as the number of patients screened, number of patients enrolled, dates of study milestones, etc. Additional reports for study oversight may be generated for the Medical Monitor at least annually.

##### 16.5 Study Records Retention

Study records will be maintained for at least seven years after the date of completion of the study.

##### 16.6 Protocol Deviations

A protocol deviation is any noncompliance with the clinical study protocol and Good Clinical Practice requirements. The noncompliance may be on the part of the subject, the investigator, or study staff. As a result of deviations, corrective actions are to be developed by the study staff and implemented promptly.

These practices are consistent with investigator and sponsor obligations in ICH E6:

- Compliance with Protocol, Sections 4.5.1, 4.5.2, 4.5.3, and 4.5.4.
- Quality Assurance and Quality Control, Section 5.1.1
- Noncompliance, Sections 5.20.1 and 5.20.2.

All deviations from the protocol must be addressed in study subject source documents and promptly reported to NIDCR and the local IRB, according to their requirements.

#### 17 PUBLICATION/DATA SHARING POLICY

This study will comply with the *NIH Public Access Policy*, which ensures that the public has access to the published results of NIH funded research. It requires scientists to submit final peer-reviewed journal manuscripts that arise from NIH funds to the digital archive PubMed Central upon acceptance for publication.

Also, the International Committee of Medical Journal Editors (ICMJE) policy requires that all clinical trials be registered in a public trials registry. In keeping with this policy, this study will be registered in *ClinicalTrials.gov*, which is sponsored by the National Library of Medicine.

Findings from this study will be presented in a timely manner at national and local scientific meetings and in the form of peer reviewed publications in scientific journals. Collaboration with others also will be welcomed. Investigators interested in collaborating or conducting an independent secondary analysis of our collected data will be asked to submit a proposal to the PI. This proposal will be evaluated by the PI and the investigative team for its scientific merit, and a decision will be made to either grant permission to the interested investigators to proceed or not. All requestors of data will need to provide evidence of IRB approval. Permission may be granted with a delay to provide a chance for the primary investigators of this study to publish their own findings. Investigators who receive approval from the PI and the investigative team to conduct a secondary analysis of the data will be granted access to a copy of the partial or full database containing only coded information. The cost incurred for the retrieval and any re-coding or transformations requested would have to be covered by the investigator requesting the use of the data.

#### 18 PROTOCOL REVISION HISTORY

| Version Date on Protocol | Version | Changes to Protocol |
|---|---|---|
| 06-Jun-2016 | 0.54 | Initial version submitted to IRB; approved 29-Aug-2016 |
| 23-Aug-2017 | 1.68 | General/Overall Revisions<br>1) Updated version numbering since a previous version (v .54) of the protocol was reviewed/approved by site IRBs<br>2) Incorporated all changes to study design following DSMB review and comments. NIDCR confirmed that the study no longer requires DSMB oversight and will receive MMOR oversight.<br>3) Revised references of "scheduled surgery" to "recommended surgery" or "in need of surgery"<br>4) Replaced all instances of "trial" with "study" since NIDCR medical monitor indicated this was not a clinical trial<br>5) Corrected formatting and replaced all instances of "participants" with "subjects"<br>Protocol Summary Revisions<br>1) Revised language used to describe surgeon-rater population<br>2) Revised study schematic to capture new study design and study schedule for patient and surgeon-rater subjects; timeframe of visit 1 for patient participants from "1 week before" to "1 day before" potential surgery<br>Potential Risks and Benefits Revisions |

| | | |
|---|---|---|
| | | 1) Revised "Benefits" sections to indicate "better surgical planning options" rather than "better treatment options"<br>2) Created separate risks/benefits sections for each study group (patient subjects and surgeon-rater subjects) |
| 16-Nov-2017 | 2.1 | 1) Changed the number of participating surgeons for "n=8" to "n= up to 10"<br>2) Increased the number of Craniofacial centers from "four" to "five" – Tufts Medical Center (TMC) being the fifth referring site<br>3) Screening Consent process was eliminated as it is no longer mandated by the Tufts IRB at participating Boston sites |
| 10-Jan-2018 | 3.0 | 1. Changed number of craniofacial centers from 5 to 6 with the addition of Shriners Hospital for Children (SHC-BOS)<br>2. Updated language on recruitment across sites and assignments of surgeon-raters to patients' data for review |

#### 19 LITERATURE REFERENCES

12. Lee, J.Y., Han, Q., Trotman, C.-A. Three-dimensional facial imaging: accuracy and considerations for clinical applications in orthodontics. *Angle Orthod.* 2004 Oct; 74(5):587-93. PMID: 15529491.
13. Faraway, J. Modeling continuous shape change for facial animation. *Statistics and Computing.* 14: 357-363, 2004.
14. Trotman, C.-A., Faraway, J.J., Phillips, C. Visual and statistical modeling of facial movement in patients with cleft lip and palate. *Cleft Palate-Craniofac. J.* 2005 May; 42(3):245-54. PMID: 15865457, PMCID: PMC3681529.
15. Trotman, C.-A., Phillips, C., Essick, G.K., Faraway, J.J., Barlow, S.M., Losken, H.W., van Aalst, J., Rogers, L. Functional outcomes of cleft lip surgery. Part I: Study design and surgeon ratings of lip disability and the need for lip revision. *Cleft Palate-Craniofac. J.* 2007 Nov; 44(6):598-606. PMID: 18177192, PMCID: PMC3646291.
16. Trotman, C.-A., Faraway, J.J., Losken, H.W., van Aalst, J. Functional outcomes of cleft lip surgery. Part II: Quantification of nasolabial movement. *Cleft Palate-Craniofac. J.* 2007 Nov; 44(6):607-16. PMID: 18177193, PMCID: PMC3681516.
17. Trotman, C.-A., Barlow, S.M., Faraway, J.J. Functional outcomes of cleft lip surgery. Part III: Measurement of lip forces. *Cleft Palate-Craniofac. J.* 2007 Nov 44(6):617-23. PMID: 18177195, PMCID: PMC3681517.
18. Essick, G.K., Phillips, C., Trotman, C.-A. Functional outcomes of cleft lip surgery. Part IV: Between- and within-participant variables affecting lip vermillion sensory thresholds. *Cleft Palate-Craniofac. J.* 2007 Nov; 44(6):624-34. PMID: 18177194, PMCID: PMC3681524.
19. Trotman, C.-A., Phillips, C., Faraway, J.J., Hartman, T., van Aalst, J. Influence of objective three-dimensional measures and movement images on surgeon treatment planning for lip revision surgery. Cleft Palate Craniofac. J. Epub 2013 Jul. 2013 Nov;50(6):684-95. PMID: 23855676, PMCID: PMC3830636.
20. Brattstrom, V., Molsted, K., Prahl-Andersen, B., Semb, G., Shaw, W.C. The Eurocleft Study: intercenter study of treatment outcome in patients with complete cleft lip and palate. Part 2: Craniofacial form and nasolabial appearance. *Cleft Palate Craniofac. J.* 2005 Jan; 42(1):69-77. PMID: 15643918.
21. Morrant, D.G., Shaw, W.C. Use of standardized video recordings to assess cleft surgery outcome. *Cleft Palate Craniofac J.* 1996 Mar; 33(2):134–42. PMID: 8695621.
22. Bennett, M.E., Tulloch, J.F.C. Understanding orthodontic treatment satisfaction from patients' perspective: a qualitative approach. Clin. Orthod. Res. 1999 May; 2(2):53-61. PMID: 10534980.
23. Creswell, J. *Research Design: Qualitative and Quantitative Approaches.* Thousand Oaks (Calif): Sage Publications; 1994.

24. Marsh, J.L. When is enough enough? Secondary surgery for cleft lip and palate patients.
*Clin. Plast. Surg.* 1990 Jan; 17(1):37-47. PMID: 2302918.
25. Strull, W.M., Lo, B., Charles, G. Do patients want to participate in medical decision making? *JAMA.* 1984 Dec 7; 252(21):2990-4. PMID: 6502860.
26. Nelson, P.A. Qualitative Approaches in Craniofacial Research. *Cleft Palate-Craniofac. J.* Epub 2008 Oct. 2009 May; 46(3):245-51. PMID: 19642761.
27. Thomson, S. B. (2011). *Sample Size and Grounded Theory.* JOAAG, Vol. 5. No. 1
28. Strauss, R.P. and H. Broder. Directions and Issues in Psychosocial Research and Methods as Applied to Cleft-Lip and Palate and Craniofacial Anomalies. *Cleft Palate-Craniofac. J.* 1991 Apr; 28(2):150-6. PMID: 2069970.
29. Office of Behavioral and Social Sciences Research, National Institutes of Health. *Qualitative Methods in Health Research: Opportunities and Considerations in Application and Review.* Bethesda, MD. December 2001, NIH Publication No. 02-5046
30. Charmaz, K. *Constructing grounded theory : a practical guide through qualitative analysis.* London; Thousand Oaks: SAGE, 2006.
31. Burawoy, M., *Ethnography unbound : power and resistance in the modern metropolis*, ed. M. Burawoy. 1991, Berkeley: Berkeley : University of California Press.
32. Burawoy, M. *The Extended Case Method. Sociological Theory* 16(1):4-33, 1998.
33. National Science Foundation. *User friendly handbook for mixed methods evaluations.* Retrieved from *http://www.nsf.gov/pubs/1997/nsf97153/* 1997.
34. Patton, M. Q. *Qualitative evaluation and research methods* . SAGE Publications, Inc. 1990.
35. Patton, M.Q. Enhancing the quality and credibility of qualitative analysis. *Health Services Res.* 1999 Dec; 34(5 Pt2):1189-208. PMID: 10591279, PMCID: PMC1089059.
36. Crebbin, W., Beasley, S.W., Watters, D.A. Clinical decision making: how surgeons do it. *ANZ J. Surg.* Epub 2013 May. 2013 Jun 83(6):422-8. PMID: 23638720.
37. Flin, R., Youngson, G., Yule, S. How do surgeons make intraoperative decisions? *Qual. Saf. Health Care.* 2007 Jun; 16(3):235-9. PMID: 17545353, PMCID: PMC2464983
38. Landis, J.R., Sharp, T.J., Kuritz, S.J. and Koch, G.G. *Mantel-Haenszel Methods; Encyclopedia of Biostatistics.* Armitage, P., Colton, J.T., eds. Wiley & Sons, London 1998; 3:2378-2391.
39. Ananth, C.V., Preisser, J.S. Bivariate logistic regression: modeling the association of small for gestational age births in twin gestations. *Stat Med.* 1999 Aug; 18(15):2011-23. PMID: 10440883.

40. Williamson, J.M., Lipstiz, S.R, Manatunga, A.K. Modeling kappa for measuring dependent categorical agreement data. *Biometrics*. 2000 Jun; 1(2):191-202. PMID: 12933519.

### APPENDICES

#### APPENDIX A: SCHEDULE OF EVENTS

| Procedures | Patients | | | Surgeon-Raters | |
|---|---|---|---|---|---|
| | Screening | Visit 1 | Visit 2[3] | Recruitment | Intervention and Decision Visit[3] |
| Sign Consent Form | | X | | X[2] | |
| Assessment of Eligibility Criteria | X | X | | | |
| Review of Medical/Dental History | X | X | | | |
| 2D Static Images | | X | X[1] | | |
| 3D Static Images | | X | X[1] | | |
| 2D Facial Videos | | X | X[1] | | |
| 3D Dynamic Statistical Modeling Images | | X | X[1] | | |
| Assessment of Unanticipated Problems | | X | X | | |
| SAFS | | | | | X |
| IDI | | | | | X |

[1] Visit 2 will only be conducted if data collection at Visit 1 is incomplete or of poor quality.
[2] Surgeon-raters will be consented, if required by governing IRB.
[3] The activities associated with this surgeon-rater visit may span up to three days.
